# Validation of the SPiRO score for the prediction of ICU admission in patients presenting with leptospirosis in tropical Australia

**DOI:** 10.64898/2026.07.17.26358361

**Authors:** Patrick Rosengren, Simon Smith, Liam Johnston, Thomas Coombs, Nicholas Cairns, Alfred Song, Megan Staples, Anna Brischetto, Ibrahim Ishmail, Hayley Stratton, Josh Hanson

## Abstract

**Objectives:** In some resource-limited settings the case-fatality rate of severe leptospirosis can exceed 50%. Early recognition of severe disease can expedite transfer to referral centres for advanced supportive care. The entirely clinical, 3-point SPiRO score can be calculated rapidly at presentation to predict a patient’s subsequent clinical course. In its derivation study, a SPiRO score of 0 had a negative predictive value (NPV) for intensive care unit (ICU) admission of 98% (95% confidence interval (CI): 96-99). In this validation cohort we sought to confirm the clinical utility of the SPiRO score and to compare its prognostic utility with other leptospirosis-specific and general disease severity scores.

**Methods:** We examined consecutive adults presenting to high-caseload hospitals in tropical Australia with laboratory-confirmed leptospirosis between June 2016 and April 2026. The SPiRO score’s ability to predict requirement for ICU admission before hospital discharge was compared with that of the leptospirosis-specific QuickLepto score and commonly used disease severity scores, namely the SOFA, qSOFA, qSOFA-lactate, NEWS-2, qNEWS, UVA and the SIRS scores.

**Results:** ICU admission was required in 62/309 (20%) episodes of leptospirosis. The SPiRO score performed as well as – or better than – all the other scores in predicting ICU admission. The Area Under the Receiver Operating Characteristic curve for the SPiRO score was 0.83 (95% CI: 0.77-0.89); only the SOFA score had a higher value: 0.84 (0.79-0.90), although the difference was not statistically significant (p=0.08). The SPiRO score had the highest NPV for ICU admission of any of the scores: 95 (95% CI: 91-97)%.

**Conclusions:** The SPiRO score can be calculated easily at the bedside at presentation to expedite the recognition of patients with leptospirosis who are most likely to deteriorate. In resource-limited settings this entirely clinical score can also help reduce unnecessary escalation of care, optimising the use of finite health resources.

## Introduction

Leptospirosis is a life-threatening zoonotic infection caused by pathogenic spirochetes of the genus *Leptospira* [1]. There are an estimated 1 million cases of leptospirosis annually resulting in ∼60,000 deaths and the loss of 2.9 million disability-adjusted life years [2, 3]. Leptospirosis is frequently a mild, self-limiting condition, however up to 15% of cases can develop rapidly progressive, severe disease [1]. In some resource-limited settings the case fatality rate of individuals with leptospirosis requiring intensive care unit (ICU) admission can exceed 50% [4]. Even in well-resourced locations, young and otherwise healthy individuals can require vasopressor support, mechanical ventilation, renal replacement therapy (RRT) and extracorporeal membrane oxygenation (ECMO) to survive their infection [5–7].

Leptospirosis has a global distribution, but the incidence is highest in rural and remote regions in tropical and subtropical low-and middle-income countries (LMIC) where humans are more likely to encounter leptospires in the environment [1, 2]. In these locations, limited diagnostic resources can hinder the recognition of severe leptospirosis, delaying the delivery of optimal therapy and the timely transfer to referral centres for advanced critical care support [1, 8].

There has therefore been interest in the development of simple prediction scores which might be able to promptly identify the patients with leptospirosis who are at greatest risk of deterioration. The entirely clinical SPiRO score was devised in tropical Australia and can be calculated at the bedside by even junior health care workers at the time of admission [9]. In the score’s derivation study if an individual at initial presentation had a systolic blood pressure >100 mmHg, normal auscultatory findings on respiratory examination and an absence of oliguria, the negative predictive value (NPV) for subsequent ICU admission was 98% [9]. Another prediction score, QuickLepto, developed in Brazil, also had prognostic utility in its derivation study although the application of this more complex score is more challenging as it uses five variables and requires laboratory support for its determination [10].

But it is important to highlight that both the SPiRO and QuickLepto scores were derived in retrospective studies of individuals with laboratory-confirmed leptospirosis [9, 10]. Even in endemic settings, leptospirosis is notoriously challenging to differentiate from other diseases, and a definite diagnosis is frequently delayed [11–13]. This potentially limits the practical applicability of leptospirosis-specific scores which have not been shown to be superior to standard patient assessment tools such as the National Early Warning Score 2 (NEWS-2) and the SOFA (Sequential Organ Failure Assessment) score in identifying individuals with severe disease [14, 15]. These widely used, validated assessment tools use the vital signs and simple laboratory tests to identify patients who require closer monitoring or escalation of care whatever the explanation for their presentation. As individuals with life-threatening leptospirosis often have fever, hypotension, tachypnoea, tachycardia and derangement of simple laboratory indices, it might be anticipated that these general disease severity scores will also help identify high-risk patients with leptospirosis, without the need for a confirmed diagnosis [16].

Far North Queensland (FNQ), a 380,000 km^2^ region of tropical Australia has the highest incidence of leptospirosis in the country [17]. Deaths are very uncommon in the region’s well-resourced health system but almost 80% of local cases present in rural or remote locations where there is less access to the laboratory support that may help identify the ∼10% of patients who need urgent transfer to a referral centre for ICU support [9, 18]. As severe leptospirosis can evolve rapidly, early recognition and transfer is crucial to reducing morbidity and mortality [19]. However, aeromedical retrieval services are necessarily limited across FNQ’s vast geographical area and these finite retrieval resources need to be used judiciously in a region where there are many competing priorities [20–22]. Therefore, from a health system perspective, identifying the individuals who do not require escalation of care is also important. In its derivation study the SPiRO score was able to do this, but could more commonly used general disease severity scores identify low-risk patients just as easily?

This study compared the ability of leptospirosis specific scores with commonly used general disease severity scores to identify individuals with severe leptospirosis at the time of their presentation to the health system. If there was no incremental value in using the leptospirosis specific scores, this would simplify the triage and assessment of patients with possible leptospirosis in locations where this life-threatening infection is endemic.

## Methods

This retrospective study was performed in the FNQ region of tropical Australia (supplementary figure 1). FNQ is home to a population of approximately 290,000 people and has a thriving agricultural sector that produces 80% of Australia’s bananas and large quantities of sugar cane, industries that are closely linked to a local incidence of laboratory-confirmed leptospirosis of approximately 15/100000 [17, 23, 24]. The study included consecutive adults (individuals aged >16 years) with laboratory-confirmed leptospirosis who were managed at four FNQ hospitals with a high leptospirosis caseload: Cairns Hospital, the 531-bed referral hospital and sole ICU for the FNQ region, and 3 smaller, peripheral hospitals in Atherton, Innisfail and Tully (56, 49 and 9 inpatient beds, respectively). The study only included individuals diagnosed with leptospirosis between 1 June 2016 and 30 April 2026 to avoid including individuals from the cohort that was used to derive the SPiRO score [9]. The study period was also chosen to coincide with the region’s December-April wet season when most cases of leptospirosis are seen in the region [24].

Potential study participants were identified by interrogating the Queensland public health system’s electronic laboratory database (AUSLAB). All participants satisfied the Australian definition of laboratory-confirmed leptospirosis, meeting one or more of the following four criteria: (1) isolation of pathogenic *Leptospira* species from blood culture; (2) a single *Leptospira* microscopic agglutination titre (MAT) ≥ 400 supported by a positive enzyme-linked immunosorbent assay IgM result; (3) a fourfold or greater rise in *Leptospira* MAT between acute and convalescent phase sera obtained at least two weeks apart or (4) detection of *Leptospira* by polymerase chain reaction (PCR) [17]. Culture of *Leptospira*, PCR testing (target outer membrane protein *LipL*32) and MAT testing were performed at Australia’s Leptospirosis Reference Laboratory in Brisbane [25].

The patients’ electronic and paper medical records were reviewed to collect demographic data and to identify any comorbidities (defined in Supplementary Table 1). The patients’ symptoms, clinical signs and laboratory data were collected for each presentation to the hospital; if patients presented multiple times within one episode of illness, these data were recorded for each presentation. If a clinical symptom or sign was not documented, it was presumed to be absent. Individuals were said to have oliguria if it was documented in the medical notes in the initial medical assessment either as a symptom or an objective finding (<20ml urine/hour).

These data were used to calculate the leptospirosis-specific SPiRO and QuickLepto scores at the time of presentation [9, 10]. The relevant vital signs and laboratory values at presentation were used to calculate other disease severity scores, specifically the NEWS2 score, the quick NEWS (qNEWS) score, the SOFA score, the quick SOFA (qSOFA) score, the modified lactate-SOFA score, the SIRS score and the universal vital assessment (UVA) score (Table 1) [14, 15, 26–30]. As most patients did not have an arterial blood gas performed, it was not possible to calculate the PaO_2_/FiO_2_ ratio in everyone; the SpO_2_/FiO_2_ (measured with pulse oximetry) was therefore collected instead [31].

**Table 1.** Variables used to calculate the SPiRO, QuickLepto, SOFA, qSOFA, qSOFA-lactate, NEWS-2. SIRS and UVA scores.

| Scores | SPiRO | QuickLepto | SOFA | qSOFA | qSOFA-Lactate | NEWS-2 | qNEWS | SIRS | UVA |
| --- | --- | --- | --- | --- | --- | --- | --- | --- | --- |
| **Vital signs variables** |  |  |  |  |  |  |  |  |  |
| Blood Pressure | SBP<100 mmHg | MAP<80 mmHg | MAP<70 mmHg or on vasopressors | SBP<100 mmHg | SBP<100 mmHg | SBP | SBP |  | SBP <90mmHg |
| Respiratory rate |  |  |  | RR >22 breaths/minute | RR >22 breaths/minute | RR | RR | RR >20 or PaCO<sub>2</sub> <32 mm Hg | RR >30 breaths |
| Consciousness |  | Lethargy or impaired GCS | Reduced GCS | GCS <15 | GCS <15 | V, P or U on AVPU | V, P or U on AVPU |  | GCS <15 |
| Heart Rate |  |  |  |  |  | HR |  | HR > 90 | HR ≥ 120 |
| Temperature |  |  |  |  |  | Temperature |  | Temperature >38°C or <36°C | Temperature <36°C |
| Oxygen saturation |  |  |  |  |  | O<sub>2</sub> Saturations |  |  | O<sub>2</sub> Saturations <92% |
| **Demographics** |  |  |  |  |  |  |  |  |  |
|  |  | Age > 40 |  |  |  |  |  |  |  |
| **Signs and symptoms** |  |  |  |  |  |  |  |  |  |
|  | Oliguria | Pulmonary symptom or sign | Mechanical ventilation |  |  | Hypercapnic respiratory failure |  |  |  |
|  | Abnormal respiratory auscultation |  |  |  |  | On supplemental oxygen |  |  |  |
| **Laboratory variables** |  |  |  |  |  |  |  |  |  |
|  |  | Haematocrit <30% | Bilirubin |  | Lactate |  |  | WBC >12,000/mm<sup>3</sup>, <4,000/mm<sup>3</sup>, or >10% bands | HIV seropositive |
|  |  |  | Platelet count |  |  |  |  |  |  |
|  |  |  | Creatinine |  |  |  |  |  |  |
|  |  |  | SpO<sub>2</sub>/FiO<sub>2</sub> |  |  |  |  |  |  |

SOFA: sepsis-related organ failure assessment; qSOFA: quick sepsis-related organ failure assessment qSOFA-Lactate: lactate enhanced quick sepsis-related organ failure assessment; NEWS-2: National Early Warning Score 2; SIRS: Systemic inflammatory response syndrome; UVA: Universal vital assessment; SBP: systolic blood pressure; MAP: mean arterial pressure. RR: respiratory rate; HR: heart rate; SpO<sub>2</sub>/FiO<sub>2</sub>: oxygen saturation: inspired fraction of oxygen ratio. GCS: Glasgow coma scale; WBC: white blood cells.

The study’s primary endpoint was a requirement for ICU admission before hospital discharge. If an individual presented and was discharged from hospital, but then represented and was admitted to ICU, the patient was said to require ICU admission for the second, but not the first presentation.

## Statistical Analysis

Data were de-identified, entered into an electronic database (Microsoft Excel, version 16.0) and analysed with statistical software (Stata version 18.5). Groups were compared using the chi-squared test, Fisher’s exact test, the Wilcoxon rank-sum test, or logistic regression, where appropriate. The predictive utility of the different severity scores was compared by calculating each score as a continuous variable and comparing the area under the receiver operating characteristic (AUROC) curves. Graphs presenting the association between the different values of each the scores and a requirement for ICU admission were constructed. These graphs were reviewed to determine the optimal cut-off for the identification of patients requiring ICU care before hospital discharge. The negative and positive predictive value for ICU admission of each of the scores using this cut-off was calculated. If individuals were missing data, they were not included in analyses which evaluated those variables.

## Ethical Approval

The study was approved by the Far North Queensland Human Research Ethics Committee (HREC EX/2024/QCH/108994). As the retrospective data were de-identified and presented in an aggregated manner, the Committee waived the requirement for informed consent.

## Results

### Characteristics of the cohort

There were 306 adults who had 309 separate episodes of laboratory-confirmed leptospirosis during the study period; 278/306 (91%) individuals were male and only 32/306 (10%) had documented comorbidity. The patients’ median (interquartile range, IQR) age at the time of presentation was 32 (25-46) years. Most episodes (202/309, 65%) occurred during the local December-April wet season. A plausible occupational or recreational exposure could be identified in 257/309 (83%) episodes; this was work in the local fruit industry in 158/257 (61%).

### Diagnosis

The diagnosis of leptospirosis was established in the 309 episodes using blood PCR in 236 (76%), initial MAT serology in 15 (5%), follow up MAT serology (fourfold rise in paired specimens) in 21 (7%) and blood culture in 37 (12%). It was possible to define or to infer a serovar in 215/309 (70%) episodes; the most commonly implicated serovars were *Leptospira interrogans* serogroup Pyrogenes serovar Zanoni (66/215, 31%) and *L. interrogans* serogroup Australis serovar Australis (44/215, 20%) (Supplementary Table 2).

### Presentation for care

The patients’ median (IQR) duration of symptoms prior to their initial hospital presentation was 3 (2-5) days. The initial presentation was to a peripheral hospital in 273/309 (88%); 83/273 (30%) were subsequently transferred to a referral centre for escalation of care. There were 29/309 (9%) who represented after initial hospital discharge and 3/309 (1%) who represented a second time (Supplementary Figure 2). Individuals presenting initially to peripheral hospitals had a shorter duration of symptoms than individuals presenting initially to the referral hospital (median (IQR): 3 (2-5) days versus 4 (3-5) days, p=0.007). Individuals working in the fruit industry had a shorter duration of symptoms at presentation than individuals without this risk factor (median (IQR): 3 (2-4) days versus 4 (3-5) days, p=0.0001).

### Clinical course

Overall, 62/309 (20%) episodes of leptospirosis required ICU admission. Vasopressor support was provided in 56/62 (90%), intubation and mechanical ventilation in 14/62 (23%), RRT in 12/62 (19%) and ECMO in 2/62 (3%) (Supplementary Figure 3).

Individuals requiring ICU admission had a longer duration of symptoms before hospital presentation than those that did not require ICU admission (median (IQR): 5 (3-6) days versus 3 (2-4) days, p=0.0001). The other epidemiological, clinical and laboratory findings at the time of the 341 presentations in the 309 episodes of leptospirosis, and the association of these findings with a subsequent requirement for ICU admission are presented in supplementary tables 3-6. There were no deaths in the 309 episodes of leptospirosis.

### Performance of the disease severity scores Utility of individual components

The association between the variables used to calculate the different scores and a requirement for ICU admission are presented in Table 2. An abnormal respiratory examination, oliguria and hypotension were three of the four clinical variables most associated with a subsequent requirement for ICU admission. It was notable that ICU admission was required less frequently in individuals who presented initially to a peripheral hospital or in those in whom leptospirosis was in the initial differential diagnosis. The presence of comorbidity had only a modest effect on the requirement for ICU admission.

**Table 2.**
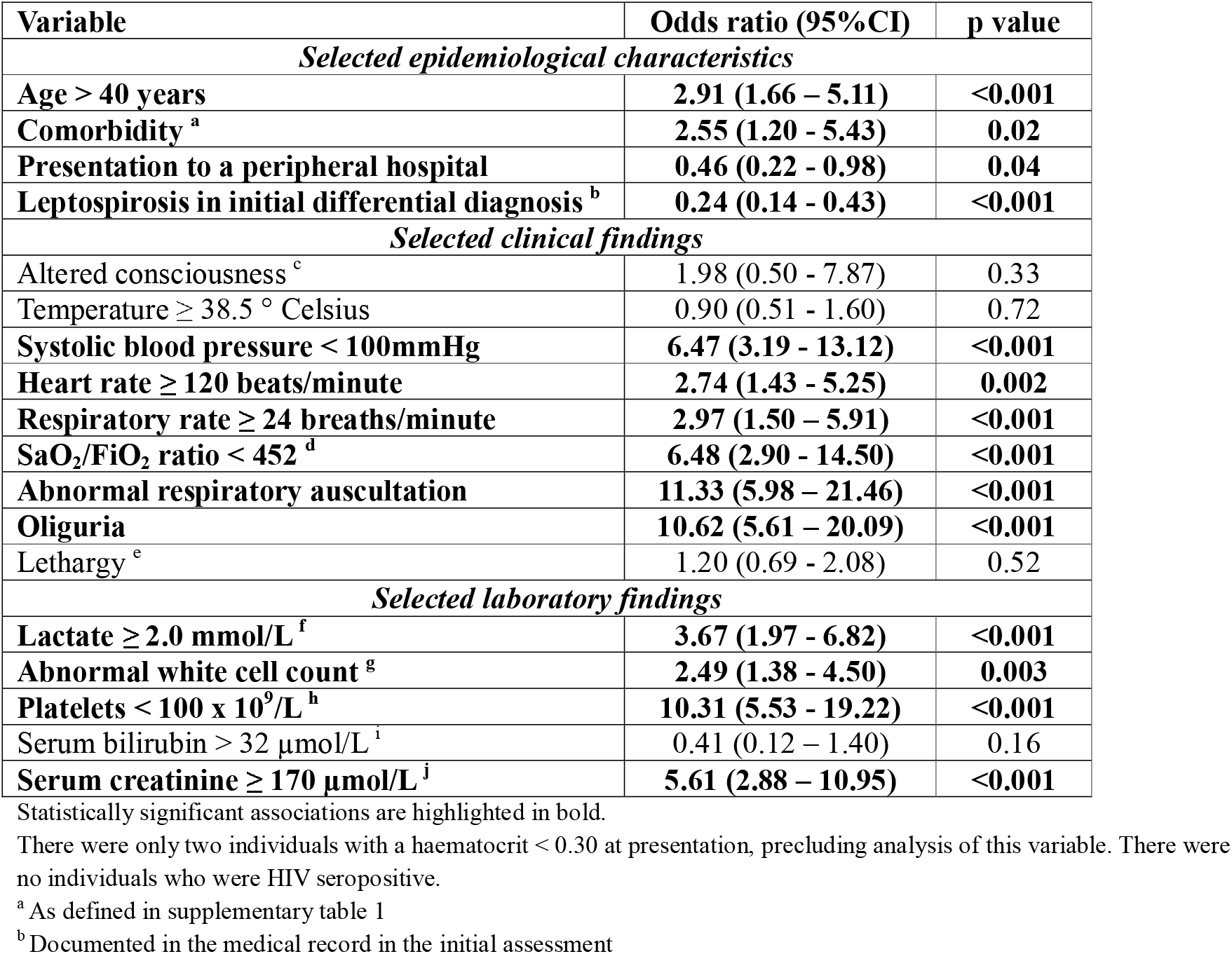
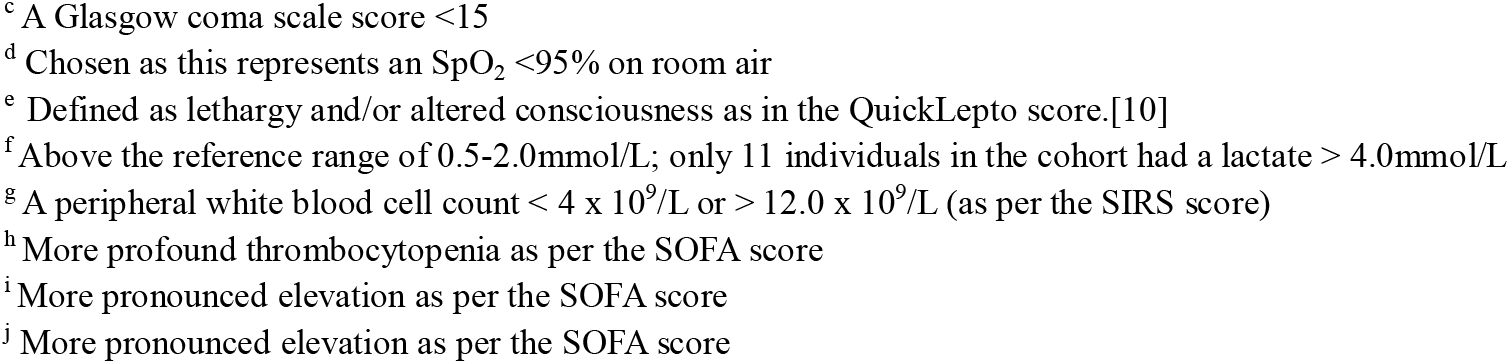
Selected variables at presentation in the 341 presentations in 309 episodes of leptospirosis and the association with a requirement for ICU before hospital discharge.

| Variable | Odds ratio (95% CI) | p value |
| --- | --- | --- |
| *Selected epidemiological characteristics* |  |  |
| Age > 40 years | **2.91 (1.66 – 5.11)** | **<0.001** |
| Comorbidity <sup>a</sup> | **2.55 (1.20 - 5.43)** | **0.02** |
| Presentation to a peripheral hospital | **0.46 (0.22 - 0.98)** | **0.04** |
| Leptospirosis in initial differential diagnosis <sup>b</sup> | **0.24 (0.14 - 0.43)** | **<0.001** |
| *Selected clinical findings* |  |  |
| Altered consciousness <sup>c</sup> | 1.98 (0.50 - 7.87) | 0.33 |
| Temperature $\geq 38.5^{\circ}$ Celsius | 0.90 (0.51 - 1.60) | 0.72 |
| Systolic blood pressure < 100mmHg | **6.47 (3.19 - 13.12)** | **<0.001** |
| Heart rate $\geq 120$ beats/minute | **2.74 (1.43 - 5.25)** | **0.002** |
| Respiratory rate $\geq 24$ breaths/minute | **2.97 (1.50 – 5.91)** | **<0.001** |
| SaO<sub>2</sub>/FiO<sub>2</sub> ratio < 452 <sup>d</sup> | **6.48 (2.90 - 14.50)** | **<0.001** |
| Abnormal respiratory auscultation | **11.33 (5.98 – 21.46)** | **<0.001** |
| Oliguria | **10.62 (5.61 – 20.09)** | **<0.001** |
| Lethargy <sup>e</sup> | 1.20 (0.69 - 2.08) | 0.52 |
| *Selected laboratory findings* |  |  |
| Lactate $\geq 2.0$ mmol/L <sup>f</sup> | **3.67 (1.97 - 6.82)** | **<0.001** |
| Abnormal white cell count <sup>g</sup> | **2.49 (1.38 - 4.50)** | **0.003** |
| Platelets < 100 x 10<sup>9</sup>/L <sup>h</sup> | **10.31 (5.53 - 19.22)** | **<0.001** |
| Serum bilirubin > 32 μmol/L <sup>i</sup> | 0.41 (0.12 – 1.40) | 0.16 |
| Serum creatinine $\geq 170$ μmol/L <sup>j</sup> | **5.61 (2.88 – 10.95)** | **<0.001** |

Statistically significant associations are highlighted in bold.
There were only two individuals with a haematocrit < 0.30 at presentation, precluding analysis of this variable. There were no individuals who were HIV seropositive.
<sup>a</sup> As defined in supplementary table 1
<sup>b</sup> Documented in the medical record in the initial assessment
<sup>c</sup> A Glasgow coma scale score <15
<sup>d</sup> Chosen as this represents an SpO<sub>2</sub> <95% on room air
<sup>e</sup> Defined as lethargy and/or altered consciousness as in the QuickLepto score.[10]
<sup>f</sup> Above the reference range of 0.5-2.0mmol/L; only 11 individuals in the cohort had a lactate > 4.0mmol/L
<sup>g</sup> A peripheral white blood cell count < 4 x 10<sup>9</sup>/L or > 12.0 x 10<sup>9</sup>/L (as per the SIRS score)

### Performance of the different disease severity scores

The SPiRO score could be calculated at presentation in 339/341 presentations. The SPIRO score performed as well as – or better than – all the other scores in predicting the requirement for ICU admission before hospital discharge (tables 3 and 4, figure 2).

**Figure 2.**
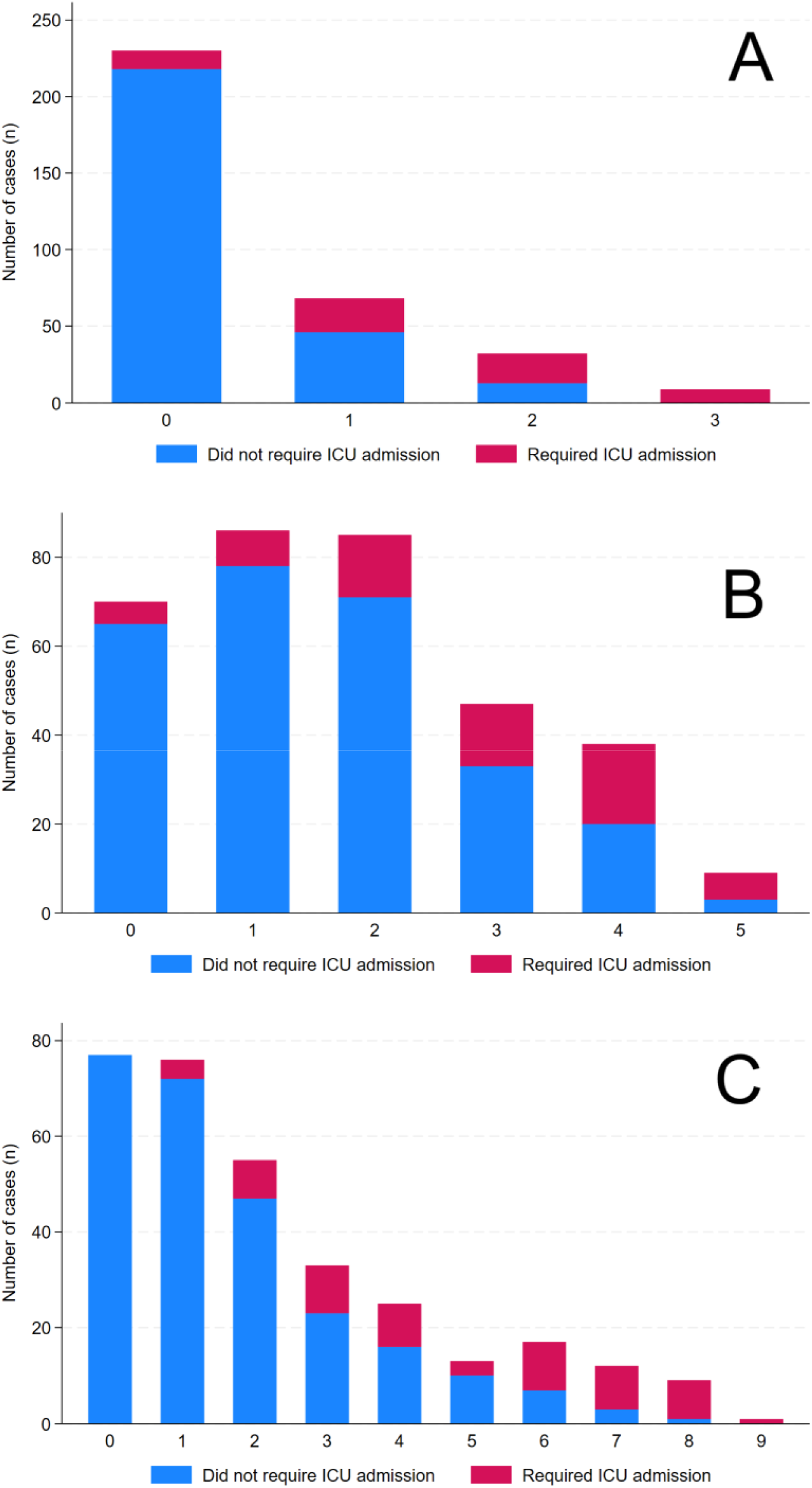
The SPiRO score (A), the QuickLepto score (B) and the SOFA score (C) in adults at presentation and the association with ICU admission before discharge. The performance of the other severity scores is presented in supplementary figures 4-12.

**Table 3.** Performance of the different severity scores in predicting ICU admission in adults.

| Score | Number of adults with sufficient data to calculate score | AUROC (95% CI) | Comparison with SPiRO score p-value |
| --- | --- | --- | --- |
| SOFA | 318 | 0.84 (0.79-0.90) | 0.08 |
| SPiRO | 339 | 0.83 (0.77-0.88) | - |
| NEWS-2 | 334 | 0.77 (0.71-0.83) | 0.08 |
| qNEWS | 334 | 0.74 (0.67-0.81) | 0.002 |
| QuickLepto | 324 | 0.74 (0.67-0.81) | 0.002 |
| qSOFA-lactate | 237 | 0.73 (0.66-0.80) | 0.02 |
| qSOFA | 334 | 0.70 (0.63-0.77) | <0.0001 |
| SIRS | 327 | 0.67 (0.60-0.74) | 0.0006 |
| UVA | 334 | 0.65 (0.58-0.72) | <0.0001 |

SOFA: Sequential Organ Failure Assessment; NEWS-2: National Early Warning Score 2; qNEWS: quick NEWS; qSOFA: quick SOFA; SIRS: Systemic Inflammatory Response Syndrome; UVA: Universal Vital Assessment; AUROC: Area Under the Receiver Operating Characteristic curve; CI: confidence interval.

**Table 4.** Comparison of the negative and positive predictive values for ICU admission of the different severity scores in adults with laboratory confirmed leptospirosis.

| Score | Number of adults with sufficient data to calculate score | Cut-off | Negative predictive value (95% CI) | Positive predictive value (95% CI) |
| --- | --- | --- | --- | --- |
| SPiRO | 339 | <1 | 95 (91-97) | 46 (36-56) |
| SOFA | 318 | <3 | 94 (90-97) | 46 (36-55) |
| QuickLepto | 324 | <2 | 92 (86-96) | 29 (22-36) |
| qNEWS2 | 334 | <2 | 91 (86-94) | 35 (27-45) |
| SIRS | 327 | <2 | 91 (84-95) | 24 (19-31) |
| NEWS-2 | 334 | <4 | 90 (86-94) | 34 (25-43) |
| qSOFA | 334 | <1 | 90 (85-93) | 35 (26-45) |
| qSOFA-lactate | 237 | <1 | 88 (81-93) | 40 (31-50) |
| UVA | 334 | <1 | 87 (83-91) | 37 (27-49) |

SOFA: Sequential Organ Failure Assessment; NEWS-2: National Early Warning Score 2; qNEWS: quick NEWS; qSOFA: quick SOFA; SIRS: Systemic Inflammatory Response Syndrome; UVA: Universal Vital Assessment; CI: confidence interval.

### Performance of the different disease severity scores in patients suspected to have leptospirosis

Leptospirosis was documented in the medical record in the initial differential diagnosis in 216/341 (63%) presentations. Leptospirosis was more commonly in the initial differential diagnosis in individuals presenting to peripheral hospitals than to referral hospitals (205/301 (68%) versus 11/40 (28%) presentations, p<0.0001) and in fruit industry workers than in those without this exposure history (149/173 (86%) versus 67/168 (40%) presentations, p<0.0001).

In an analysis restricted to the 216 individuals with leptospirosis in the initial differential diagnosis, the SPiRO score could be calculated at admission in 215/216 and performed as well as – or better than – all the other scores in predicting the requirement for ICU admission before hospital discharge (Tables 5 and 6, Figure 3).

**Figure 3.**
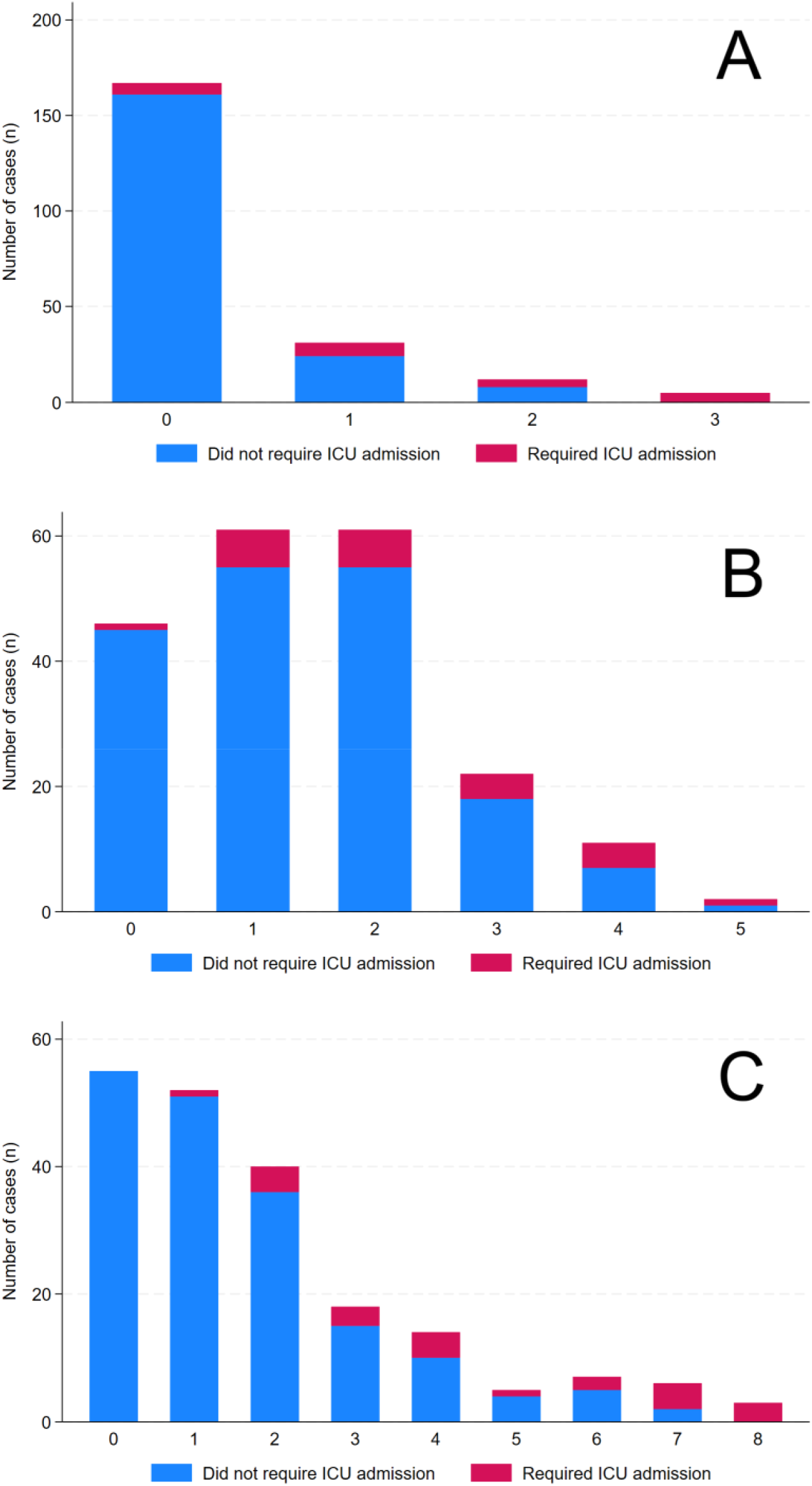
The SPiRO score (A), the QuickLepto score (B) and the SOFA score (C) in adults at presentation in whom leptospirosis was in the initial differential diagnosis and the association with ICU admission before discharge.

**Table 5.** Performance of different severity scores in predicting ICU admission in whom leptospirosis was documented in the initial differential diagnosis.

| Score | Number of adults with sufficient data to calculate score | AUROC ((95% CI) | Comparison with SPiRO score p value |
| --- | --- | --- | --- |
| SOFA | 200 | 0.88 (0.81-0.94) | 0.07 |
| SPiRO | 215 | 0.80 (0.70-0.91) | - |
| QuickLepto | 203 | 0.69 (0.58-0.81) | 0.03 |
| NEWS-2 | 212 | 0.69 (0.56-0.81) | 0.02 |
| qSOFA-lactate | 144 | 0.69 (0.57-0.81) | 0.04 |
| qNEWS | 212 | 0.67 (0.54-0.79) | 0.003 |
| qSOFA | 212 | 0.65 (0.53-0.77) | 0.0006 |
| SIRS | 206 | 0.64 (0.52-0.77) | 0.02 |
| UVA | 212 | 0.63 (0.51-0.75) | 0.002 |

SOFA: Sequential Organ Failure Assessment; NEWS-2: National Early Warning Score 2; qNEWS: quick NEWS; qSOFA: quick SOFA; SIRS: Systemic Inflammatory Response Syndrome; UVA: Universal Vital Assessment; AUROC: Area Under the Receiver Operating Characteristic curve; CI: confidence interval.

**Table 6.** Comparison of the of the different severity scores’ negative and positive predictive values for ICU admission in whom leptospirosis was documented in the initial differential diagnosis.

| Score | Number of adults with sufficient data to calculate score | Cut-off | Negative predictive value (95% CI) | Positive predictive value (95% CI) |
| --- | --- | --- | --- | --- |
| SOFA | 200 | <3 | 97 (92-99) | 32 (20-46) |
| SPiRO | 215 | <1 | 96 (92-99) | 33 (20-48) |
| QuickLepto | 203 | <2 | 94 (87-93) | 16 (9-25) |
| qNEWS2 | 212 | <2 | 93 (88-97) | 18 (10-27) |
| qSOFA | 212 | <1 | 93 (88-97) | 19 (10-31) |
| SIRS | 206 | <2 | 93 (85-98) | 13 (8-20) |
| qSOFA-lactate | 144 | <1 | 92 (84-97) | 24 (15-36) |
| UVA | 212 | <1 | 92 (87-96) | 20 (9-34) |
| NEWS-2 | 212 | <4 | 92 (86-96) | 15 (8-26) |

SOFA: Sequential Organ Failure Assessment; NEWS-2: National Early Warning Score 2; qNEWS: quick NEWS; qSOFA: quick SOFA; SIRS: Systemic Inflammatory Response Syndrome; UVA: Universal Vital Assessment; CI: confidence interval.

## Discussion

The simple 3-point SPiRO score, when calculated at presentation, was able to predict a requirement for ICU admission before hospital discharge better than, or as well as, all the other scores that were reviewed in this study. Hypotension, abnormal findings on respiratory auscultation and oliguria, the three elements of the SPiRO score, each had significant prognostic utility in this cohort and potentially also identify the cardiovascular, respiratory and renal complications that are most consistently associated with life-threatening leptospirosis [5, 6, 16, 32]. Recognition of these complications can inform immediate clinical management strategies, particularly the prompt use of vasopressors, cautious fluid resuscitation and early referral for RRT that have been linked to better outcomes [7, 33, 34].

The SPiRO score is an entirely clinical score, which is simple to calculate and can be determined at the bedside by any clinician. These characteristics make it well suited for use in rural and remote resource-limited limited settings where leptospirosis is more common and where there is often less access to laboratory support [2]. This validation series replicates the findings of the original derivation series and suggests that the SPiRO score can help expedite the recognition of patients with severe leptospirosis and by helping reduce unnecessary transfers, optimise the use of finite health resources [9].

The only score with comparable performance to the SPiRO score in this study was the SOFA score, however, the biochemistry and haematology values which represent three of the SOFA score’s six variables need laboratory support and then require weighted calculation to generate a final value. General disease severity scores including the NEWS-2, the qSOFA, the qSOFA-lactate, the SIRS and the UVA score were not as helpful in identifying patients with severe leptospirosis in this cohort. While patients presenting with leptospirosis frequently have abnormal vital signs and laboratory investigations [19, 35], important components of these general disease severity scores may be normal in leptospirosis. Impaired consciousness is rare in patients with leptospirosis (fewer than 3% of presentations in this cohort) and an elevated lactate is seen less commonly: almost three-quarters of those who had a serum lactate measured in this cohort had a normal value [36, 37]. There were no patients in our cohort who were HIV seropositive, an important component of the UVA score that was developed in sub-Saharan Africa for predicting mortality in resource-limited settings [30].

The SPiRO score was also superior to the QuickLepto score in identifying patients at risk of severe disease. This is likely to be explained by the fact that two of the QuickLepto score’s five variables had no prognostic utility in our cohort: the presence of lethargy and/or altered consciousness had no association with severe disease, and the haematocrit was less than 0.30 in only two presentations. Age over 40 is weighted in the QuickLepto score, but almost two-thirds of our cohort were aged less than 40 years, a reflection of the high incidence in young, healthy individuals working in the local banana and sugar industries.

Some authors have suggested that as prediction scores like SPiRO and QuickLepto were derived in cohorts of patients with confirmed leptospirosis, they may have less real-world utility as the diagnosis of leptospirosis may not ever be confirmed in regions where the disease is endemic [1]. However, this study demonstrates that in endemic regions like FNQ the diagnosis of leptospirosis is frequently considered by the clinicians who first assess the patient and therefore disease specific scores like SPiRO may have clinical utility even before the diagnosis is confirmed. The NPV of a SPIRO score of 0 in patients in whom leptospirosis was in the initial differential diagnosis was 96% (95% CI 92-99) compared with 95% (95% CI 91-97) in the overall cohort.

It should be noted that while the SPiRO score was very helpful in identifying the low-risk patient, the PPV of a score > 0 for ICU admission was only 46% (95% CI 36-57). However, a case can be made that a patient with a 46% chance of ICU admission is a patient in whom transfer to a referral centre should be strongly considered, or, at the very least, who should be monitored more closely.

The entirely clinical nature of the SPiRO score is one of its virtues, but it should be emphasised that this does not obviate the need to perform the laboratory testing and relevant imaging that will enhance patient care. Other diagnoses need exclusion, electrolytes may need correction, myocardial injury may need quantification, blood products may need transfusion and criteria for RRT may need to be established. It should also be highlighted that any prediction score should not replace clinical judgement; other factors or clinical findings might necessitate a requirement for escalation of care [38].

Our study has several limitations. Data were collected retrospectively which necessarily resulted in missing data, although this was mitigated by use of an electronic medical record where documentation was generally excellent. Oliguria, one of the key variables of the SPiRO score, cannot be demonstrated objectively immediately, although a focussed history may elicit the presence of this symptom and in our cohort the presence of oliguria could be defined promptly, and certainly more rapidly than the time it would take for results of laboratory investigations to be returned to the requesting clinician. In the SPiRO score’s derivation series the endpoint was severe disease (defined as ICU admission or pulmonary haemorrhage), while in this validation series we used the endpoint of ICU admission alone. We chose the modified endpoint in this study as we felt that ICU admission was a harder endpoint which necessitated a change in the location of care. There is significant geographic heterogeneity in the prevalence of different serovars which may contribute to a variation in the clinical phenotype of leptospirosis that is seen in different parts of the world [39–41]. This may limit the generalisability of our findings, although there are more similarities than differences in the clinical phenotype of severe leptospirosis in different geographic locations, with pulmonary and renal involvement and hypotension the most common life-threatening manifestations of severe disease [16]. Importantly most of the individuals in our cohort were young and otherwise healthy, which may have underestimated a contribution of age and comorbidity to a requirement for ICU [5, 10]. It is also important to highlight that our patients were managed in Australia’s well-resourced, universal health system. Patients presented relatively early in their disease course (after a median of 3 days versus a median of 7 days in the derivation study of QuickLepto [10]) and were managed by clinicians familiar with leptospirosis, which likely explains the excellent outcomes. Vital signs-based scores can be calculated repeatedly to follow a patient’s progress, which is important as the failure of vital signs to normalise is more informative than their initial values [42]. Sequential calculation of the SPiRO score has not been examined, although, as it examines the key organ systems that are affected in life-threatening leptospirosis there is every likelihood that it would also be useful.

Acknowledging these limitations, this validation series confirms that the simple SPiRO score can be calculated at presentation to help identify the high-risk patient with leptospirosis, inform their optimal care and where necessary, expedite their transfer to a referral centre. It is likely to have even more utility with the evolution of reliable point of care testing for the disease [43]. Future prospective studies would examine its application in clinical practice, how best to promptly assess oliguria and the role of sequential measurement to identify the deteriorating patient.

## Conclusions

In this validation series, the SPiRO score performed as well – or better than – all the other severity scores assessed in this study in identifying the high-risk patient with suspected leptospirosis. The score is entirely clinical and is simple to calculate making it well suited for use in the rural and resource-limited settings where, globally, most cases of leptospirosis are seen. It has the potential to enhance patient management by not only recognising individuals at higher risk of deterioration, but also identifying – and prompting therapy for – the cardiovascular, respiratory and renal complications that are most associated with life-threatening disease.

## Data availability statement

Data cannot be shared publicly because of the Queensland Public Health Act 2005. Data are available from the Far North Queensland Human Research Ethics Committee (contact via) for researchers who meet the criteria for access to confidential data.

## Supplementary material

**Supplementary Figure 1.**
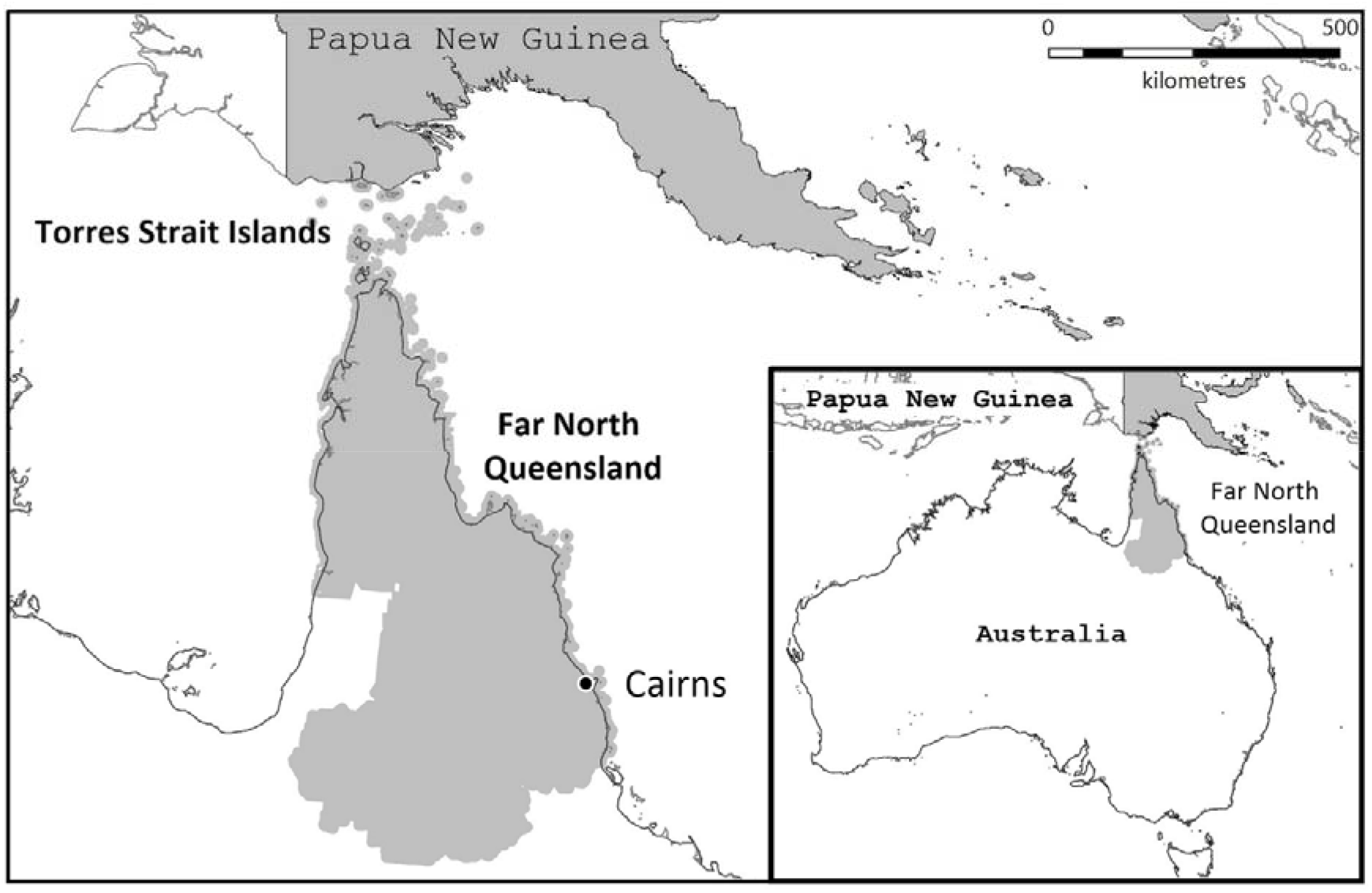
Map of Far North Queensland, Australia, showing location of the current study.

**Suppementary Figure 2.**
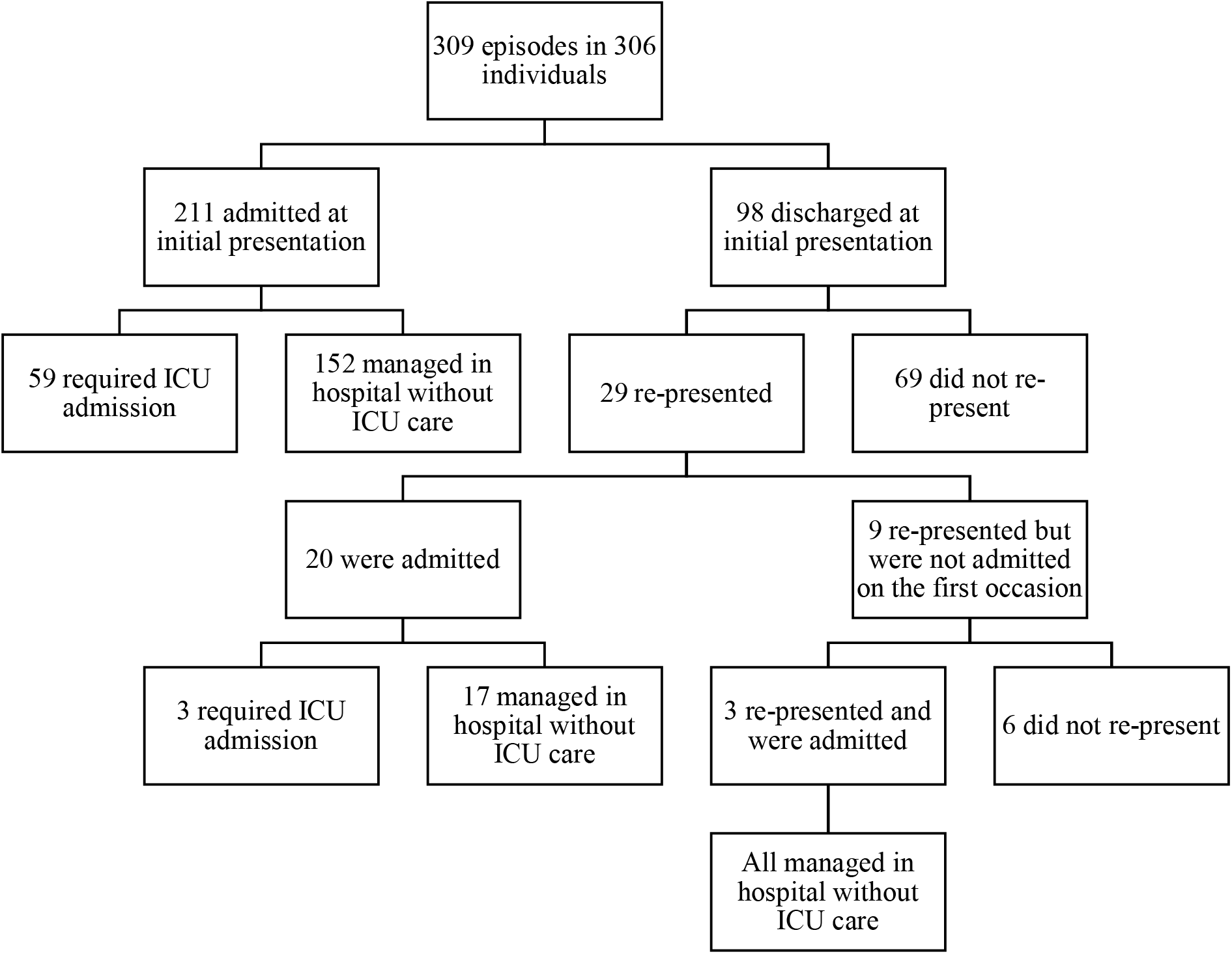
Flow diagram showing presentation and clinical course of individuals with laboratory confirmed leptospirosis in the study

**Supplementary Figure 3.**
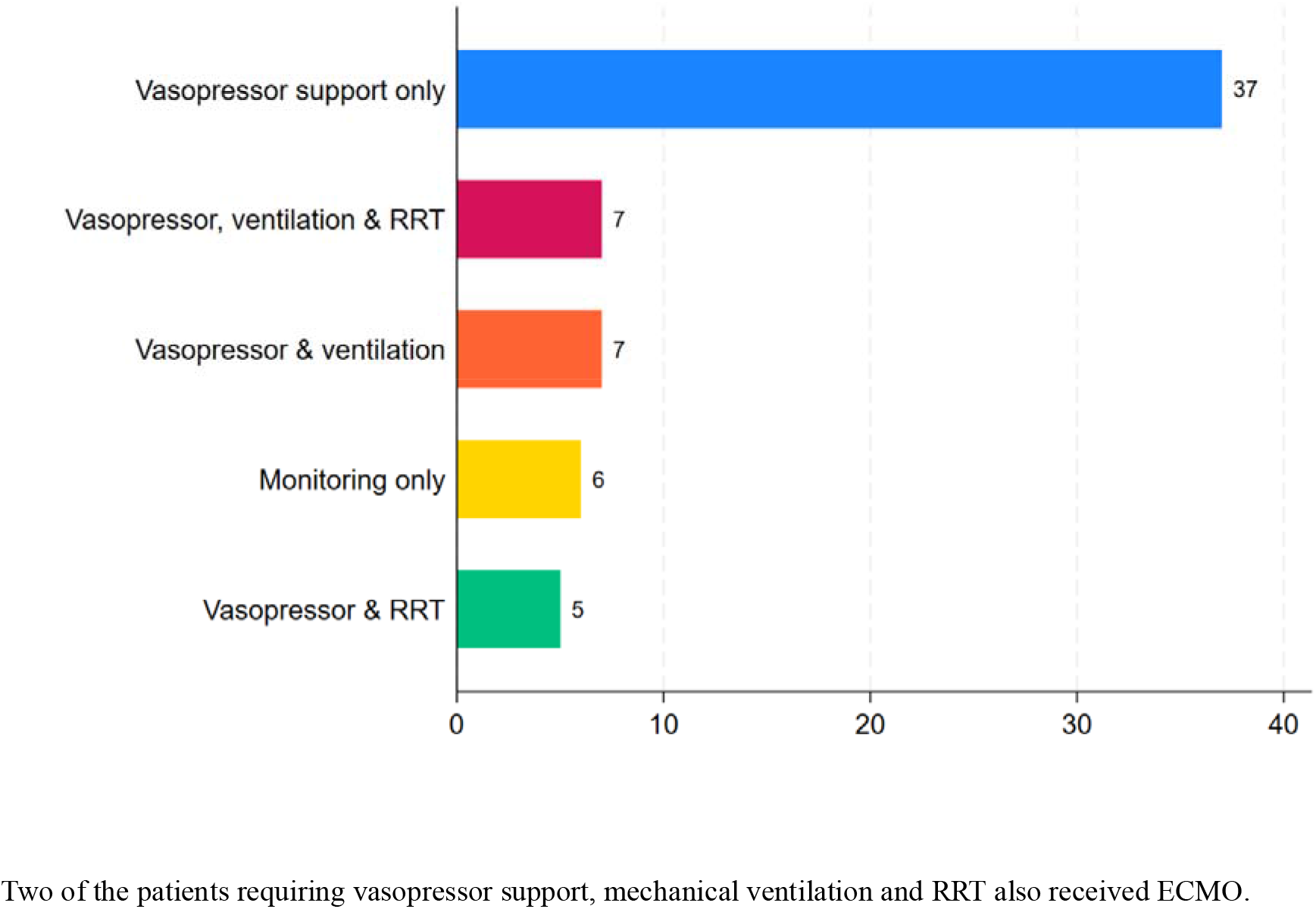
Bar chart describing the support provided to the 62 individuals who were admitted to the ICU during the study period. Two of the patients requiring vasopressor support, mechanical ventilation and RRT also received ECMO.

**Supplementary Figure 4.**
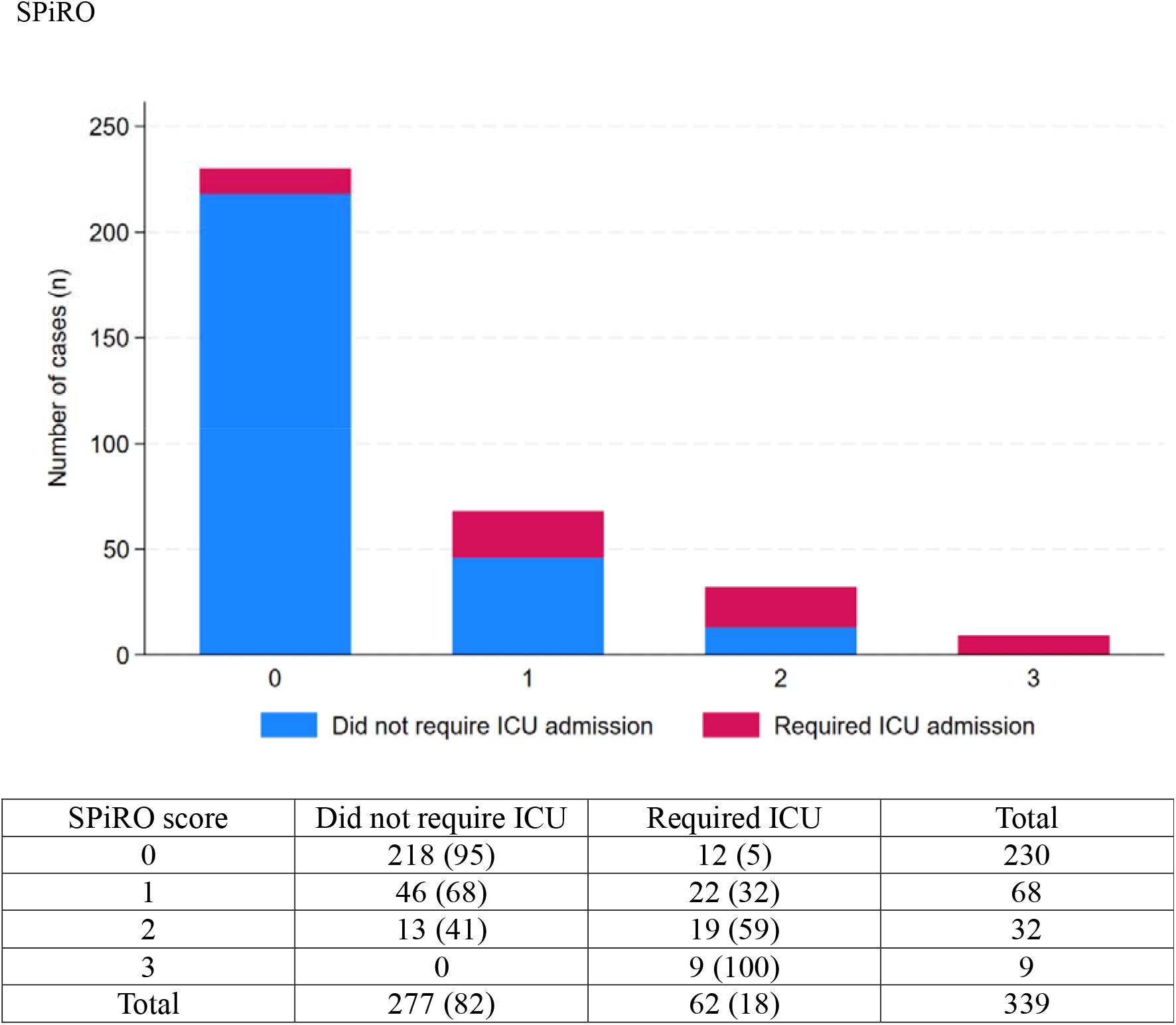
The SPIRO score in adults with leptospirosis at presentation and the association with subsequent ICU admission

**Supplementary Figure 5.**
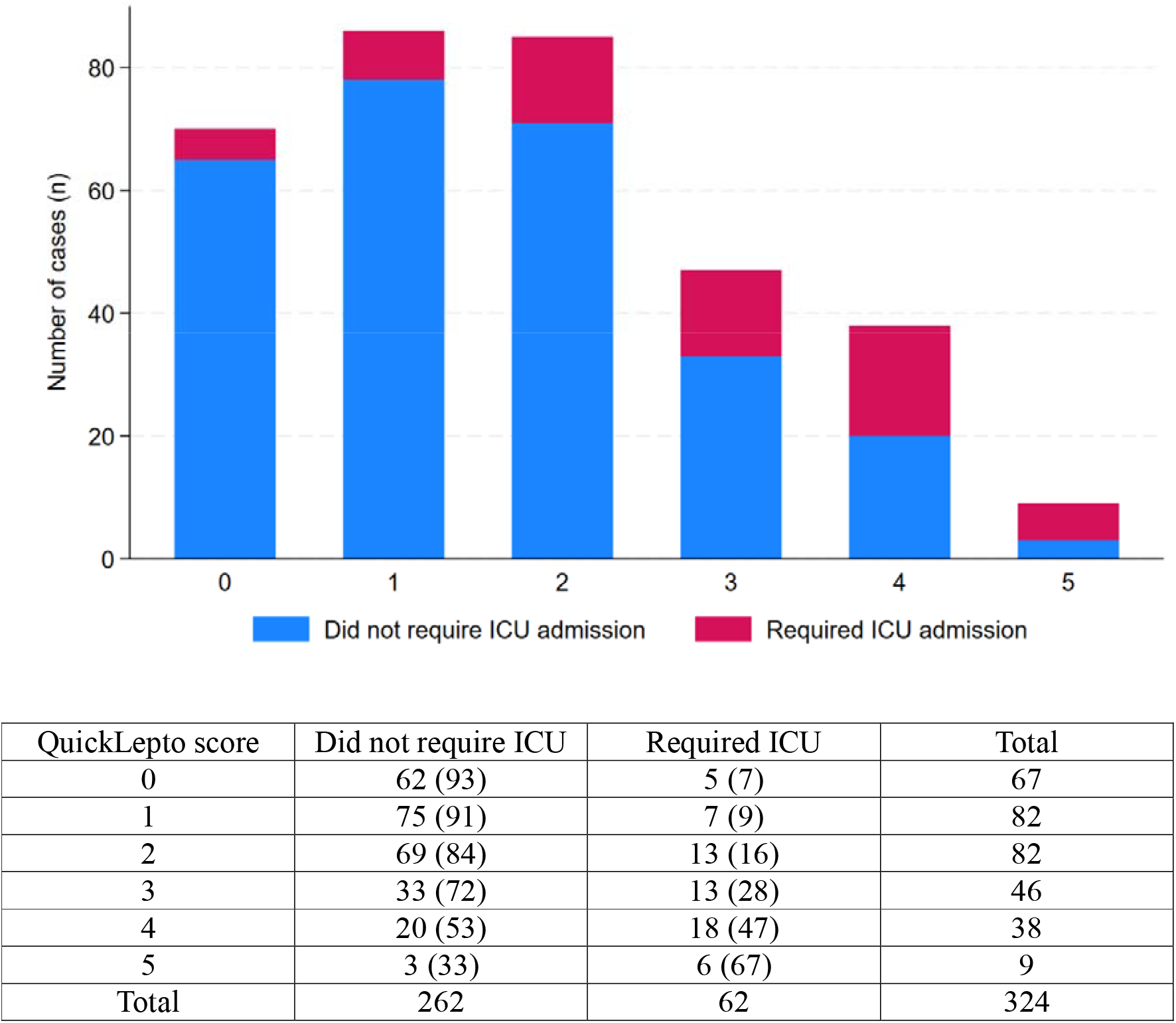
The QuickLepto score in adults with leptospirosis at presentation and the association with subsequent ICU admission

**Supplementary Figure 6.**
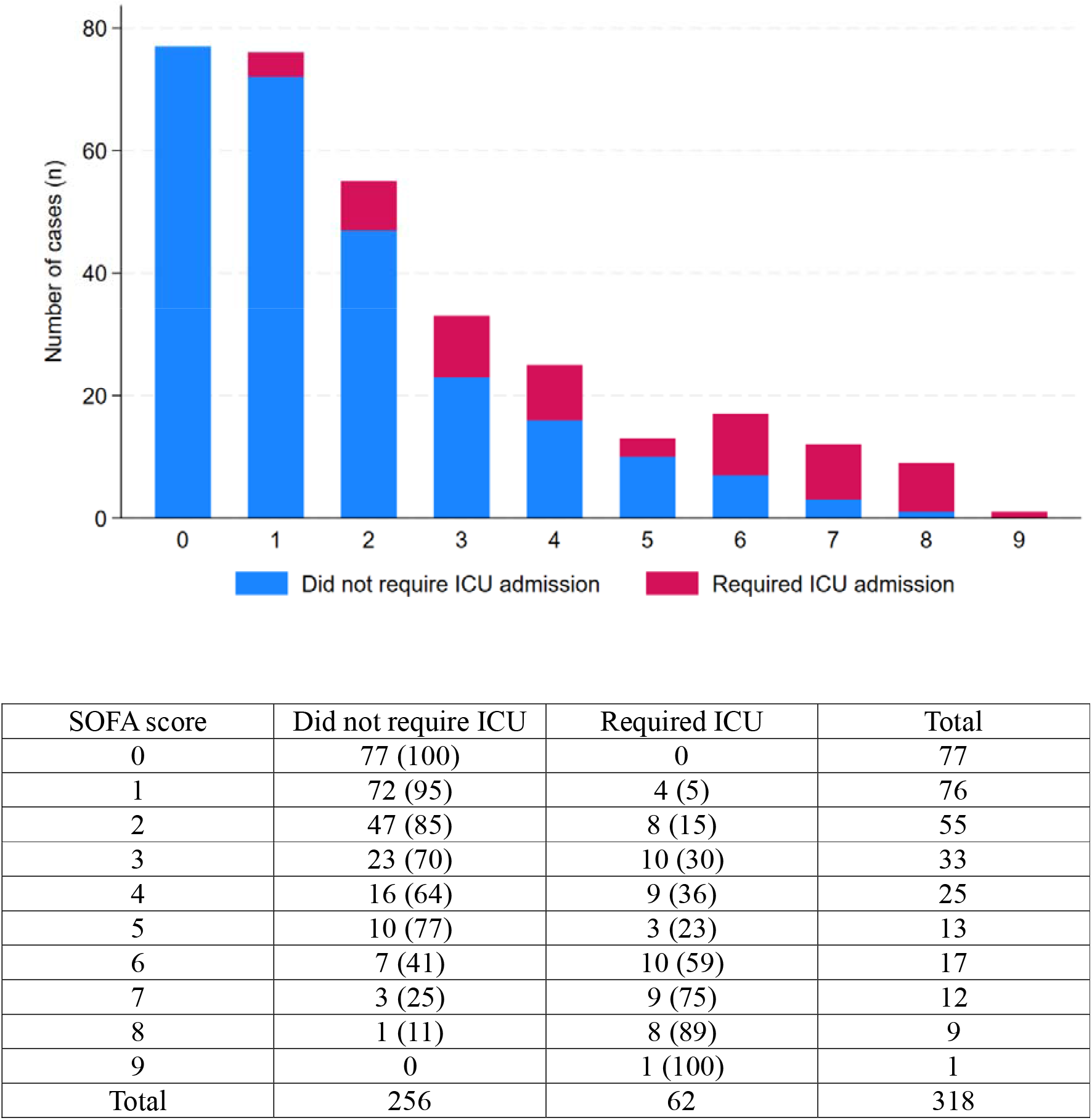
The SOFA score in adults with leptospirosis at presentation and the association with subsequent ICU admission

**Supplementary Figure 7.**
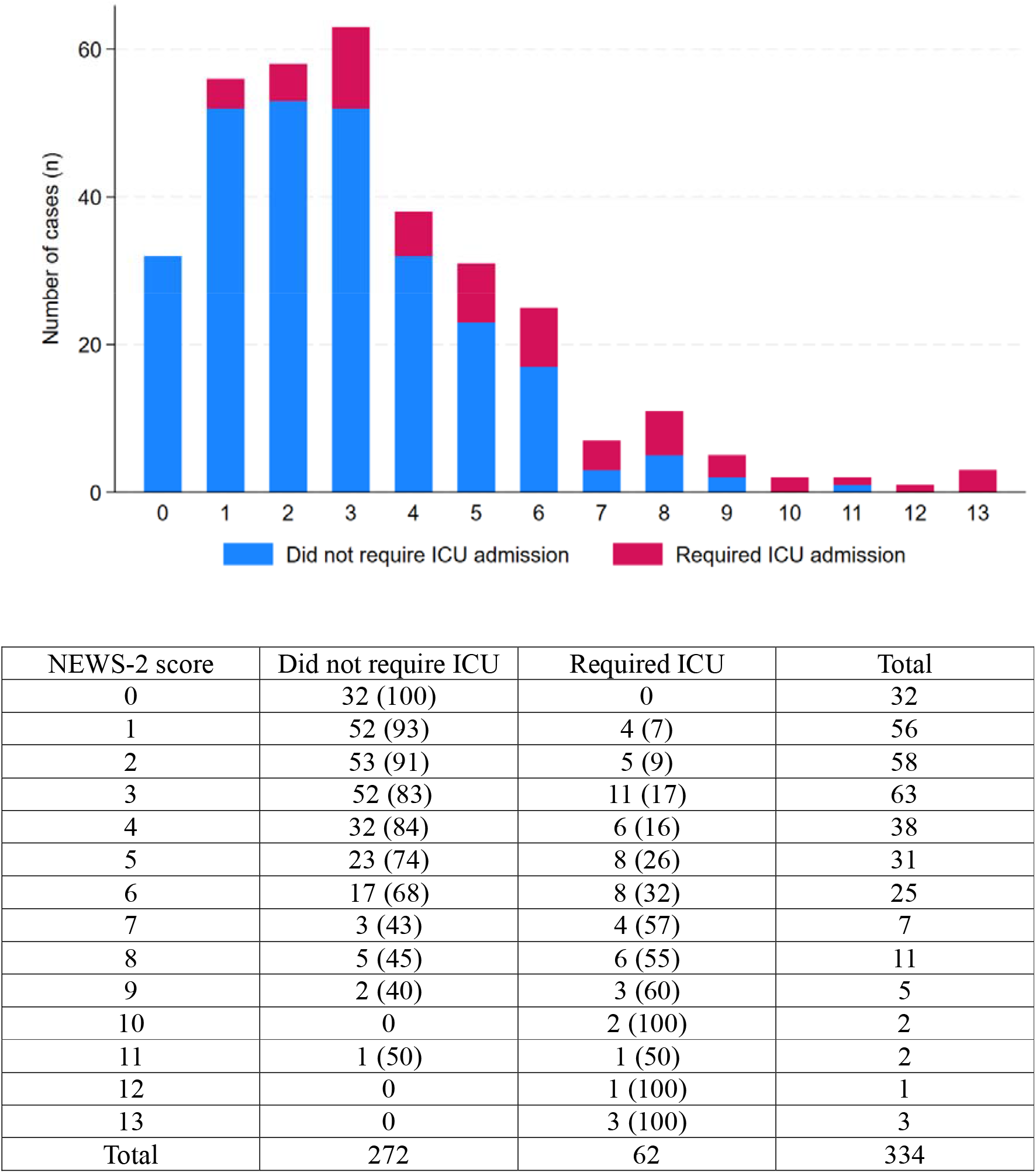
The NEWS-2 score in adults with leptospirosis at presentation and the association with subsequent ICU admission

**Supplementary Figure 8.**
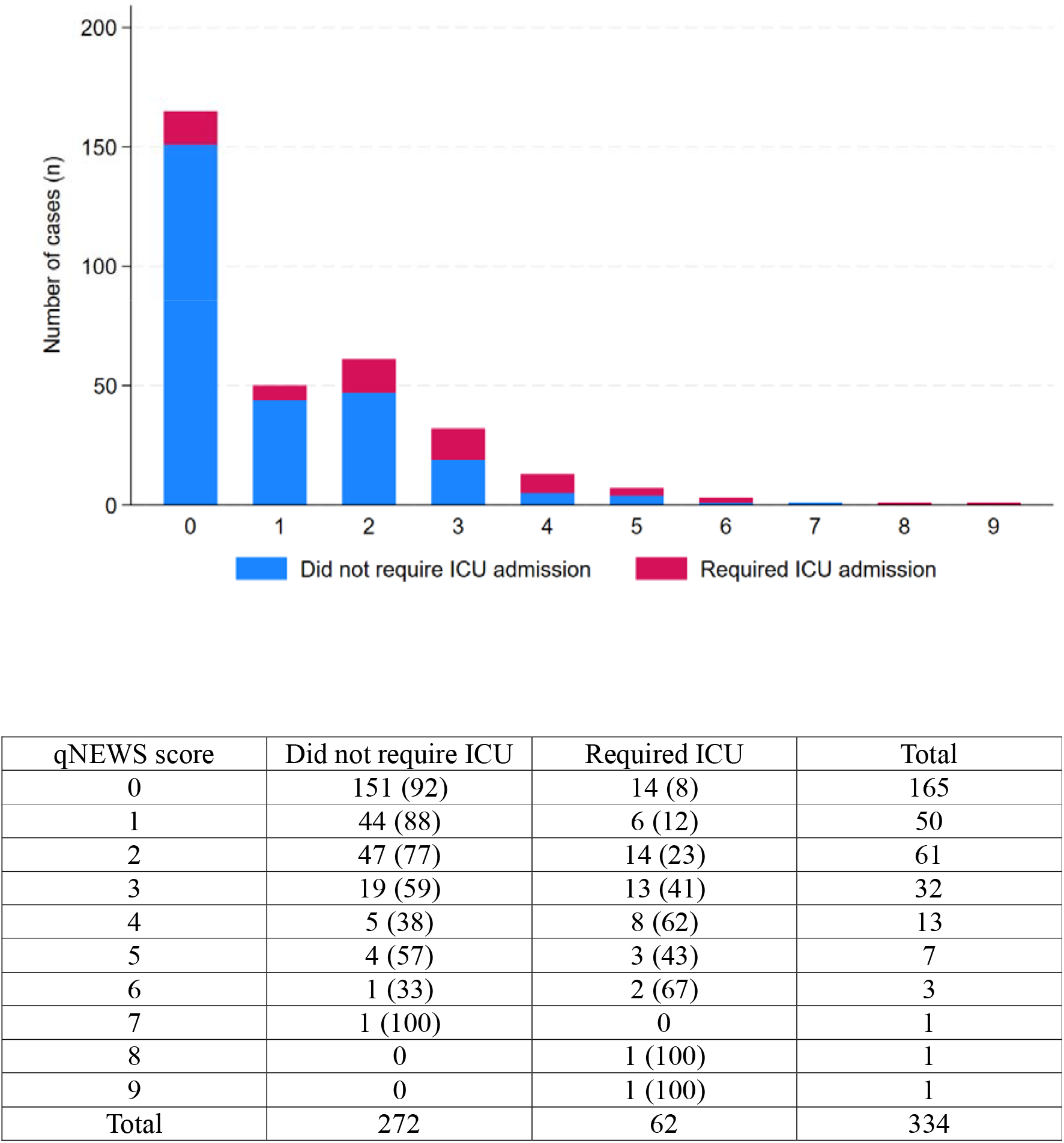
The qNEWS score in adults with leptospirosis at presentation and the association with subsequent ICU admission

**Supplementary Figure 9.**
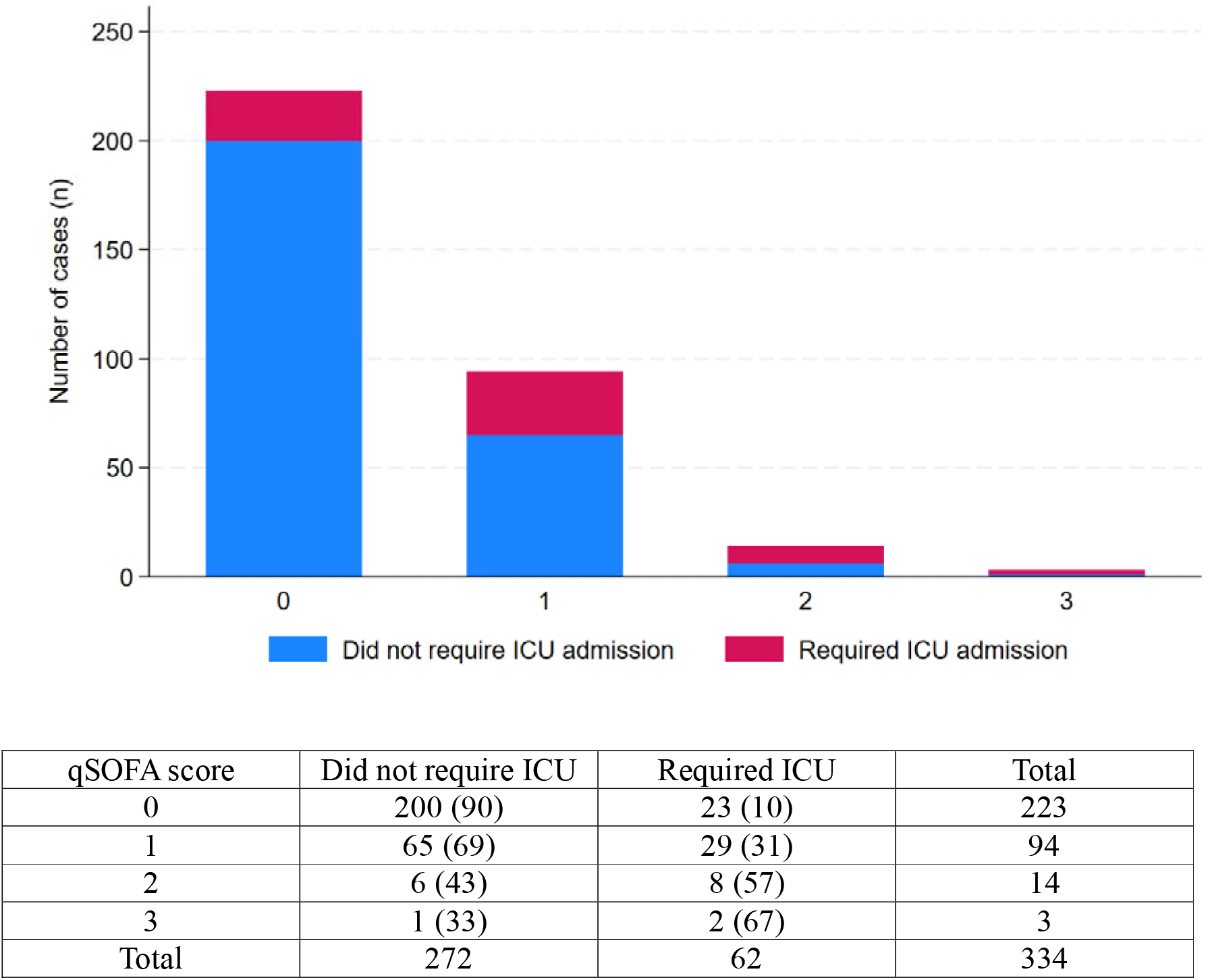
The qSOFA score in adults with leptospirosis at presentation and the association with subsequent ICU admission

**Supplementary Figure 10.**
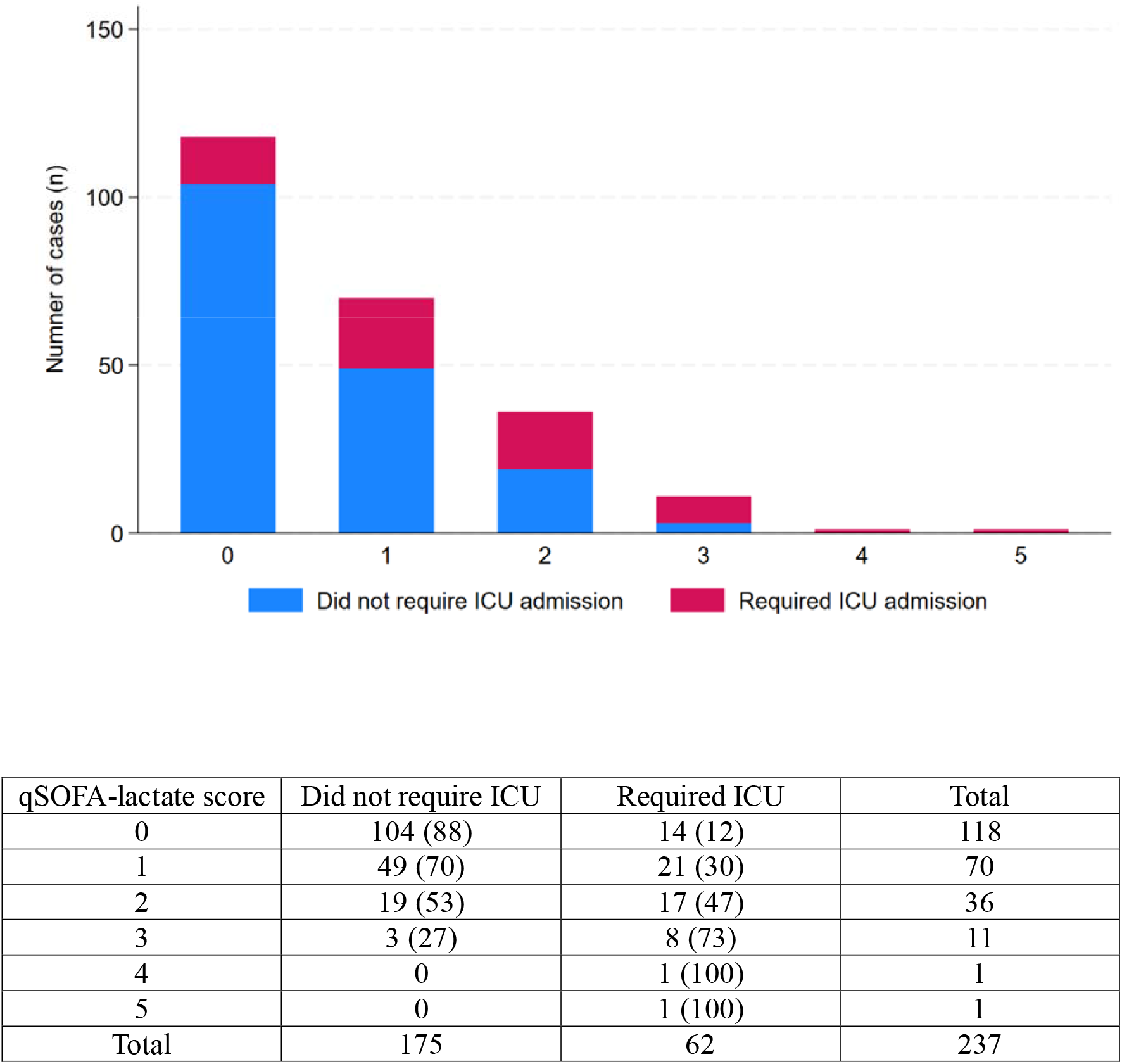
The qSOFA-lactate score in adults with leptospirosis at presentation and the association with subsequent ICU admission

**Supplementary Figure 11.**
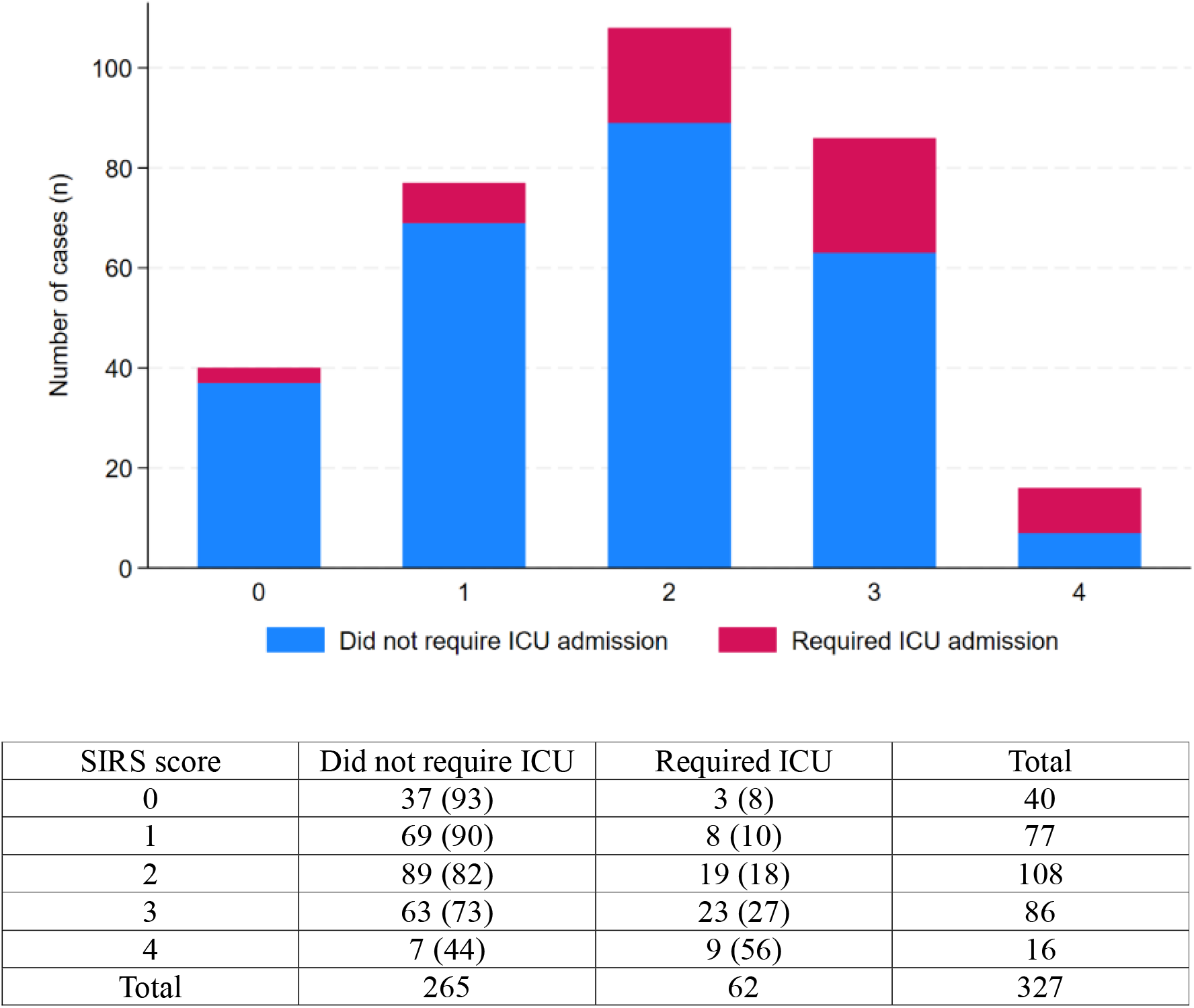
The SIRS score in adults with leptospirosis at presentation and the association with subsequent ICU admission

**Supplementary Figure 12.**
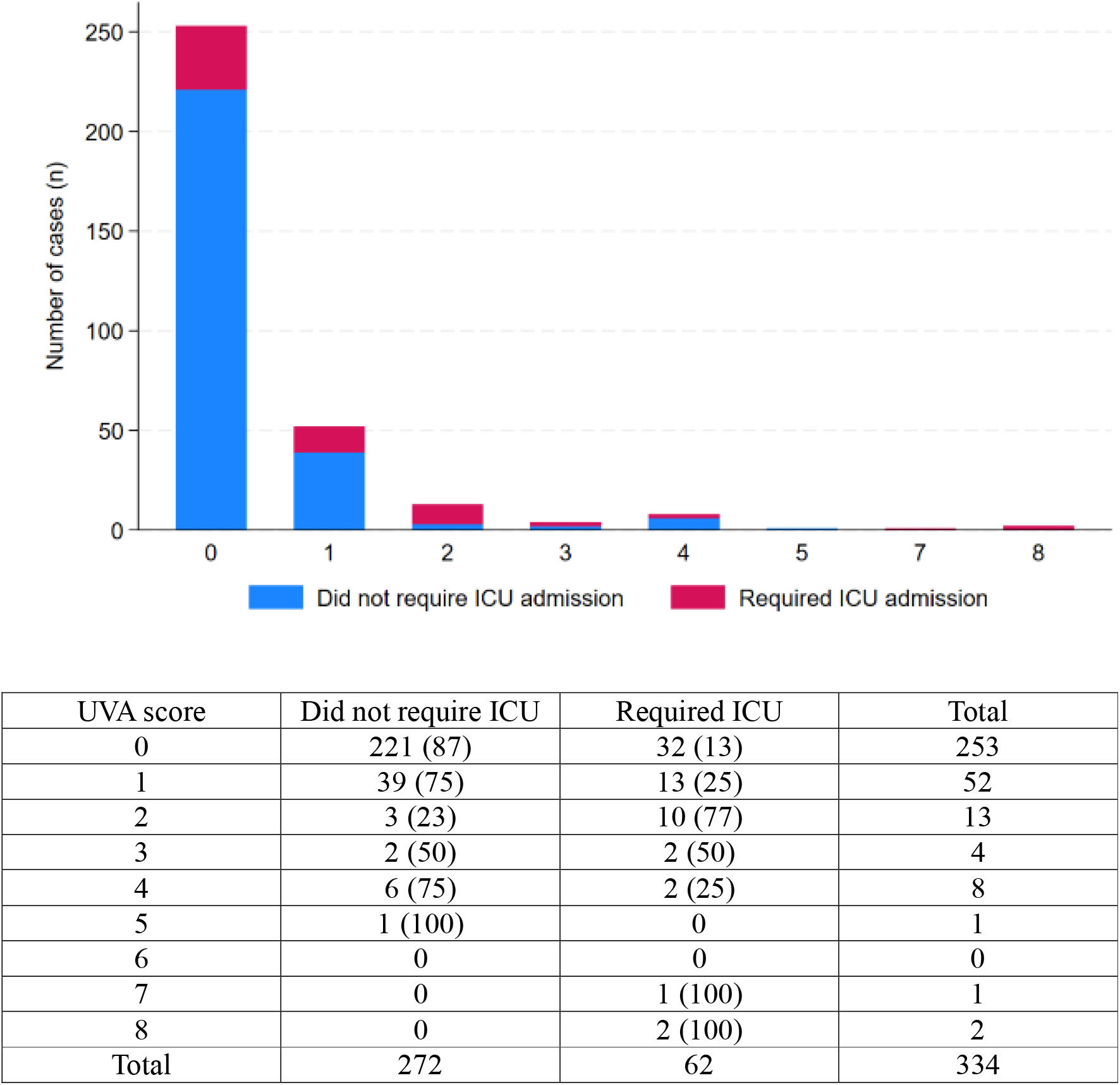
The UVA score in adults with leptospirosis at presentation and the association with subsequent ICU admission

---

**Supplementary Table 1.** Definitions used for comorbidities in the cohort.

| Comorbidity <sup>a</sup> |  | Definition used |
| --- | --- | --- |
| If documented in patient's medical history recorded by the admitting clinician | Diabetes mellitus | Documented in history, HbA1c > 6.5%, or on therapy for diabetes mellitus |
|  | Chronic heart failure | Documented in history, on heart failure therapy, or a history of an echocardiogram demonstrating LVEF < 50% |
|  | Ischaemic heart disease | Documented in history as having had a myocardial infarction or coronary artery bypass graft |
|  | Chronic kidney disease | Documented in history or if baseline eGFR <90 |
|  | Chronic lung disease | Documented in history; included interstitial lung disease, chronic obstructive pulmonary disease, bronchiectasis, cystic fibrosis, asthma or any other disease requiring therapy |
|  | Chronic liver disease | Documented as having cirrhosis (notes or imaging) |
|  | Active malignancy | Documented in history |
|  | Autoimmune condition | Documented in history |
|  | Immunosuppression | If patient was taking any medications to suppress their immune system including regular corticosteroids, other immunosuppressants or immunomodulatory therapy |
|  | Hazardous alcohol intake | Regular intake of >10 standard drinks per week or >4 standard drinks in one day at least once/month |
|  | Smoker | Smoking tobacco on a regular basis |

HbA1c: glycosylated haemoglobin; LVEF: left ventricular ejection fraction; eGFR: estimated glomerular filtration rate; CKD: chronic kidney disease.
<sup>a</sup> Categorical variable

**Supplementary Table 2.** Serovars of the 215 episodes of leptospirosis in which a serovar was identified or inferred.

| **Serovar** | **n (%)** |
| --- | --- |
| Zanoni | 66 (31%) |
| Australis | 45 (21%) |
| Arborea | 42 (20%) |
| Kremastos | 11 (5%) |
| Robinsoni | 11 (5%) |
| Topaz | 7 (3%) |
| Celledoni | 6 (3%) |
| Szwajizak | 5 (2%) |
| Copenhageni | 4 (2%) |
| Hardjo | 4 (2%) |
| Pomona | 2 (1%) |
| Bulgarica | 1 (0.5%) |
| Cynopteri | 1 (0.5%) |
| Djasiman | 1 (0.5%) |
| Javanica | 1 (0.5%) |
| Zanoni, Robinsoni <sup>a</sup> | 3 (1%) |
| Arborea, Robinsoni <sup>a</sup> | 1 (0.5%) |
| Hardjo, Bataviae <sup>a</sup> | 1 (0.5%) |
| Kremastos, Djasiman <sup>a</sup> | 1 (0.5%) |
| Topaz, Tarassovi <sup>a</sup> | 1 (0.5%) |
| Szwajizak, Medanensis, Bataviae <sup>a</sup> | 1 (0.5%) |
| Total | 215 |

<sup>a</sup> In eight cases MAT cross reactivity precluded identification of a single serovar

**Supplementary Table 3.** Selected epidemiological characteristics of the 309 episodes of leptospirosis (occurring in 306 individuals) and their association with a requirement for ICU admission during that episode.

|  | Number with data | All | Did not require ICU admission <sup>a</sup><br>n=247 | Required ICU admission <sup>a</sup><br>n=62 | p |
| --- | --- | --- | --- | --- | --- |
| **Age** | **309** | **32 (24-46)** | **30 (23-43)** | **44 (27-64)** | **0.0001** |
| Male gender | 309 | 281 (91%) | 226 (92%) | 55 (85%) | 0.47 |
| **Fruit industry worker** | **309** | **158 (51%)** | **143 (58%)** | **15 (24%)** | **<0.0001** |
| **Comorbidity <sup>b</sup>** | **309** | **32 (10%)** | **20 (8%)** | **12 (19%)** | **0.009** |

Median (interquartile range) or absolute number (%) presented as appropriate.
Statistically significant associations are highlighted in bold.
<sup>a</sup> During the episode of leptospirosis
<sup>b</sup> As defined in supplementary table 1

**Supplementary Table 4.** Symptoms at presentation in the 341 presentations in 309 episodes of leptospirosis and the association with a requirement for ICU during that admission.

|  | All presentations<br>n=341 | Did not require ICU admission <sup>a</sup><br>n=279 | Required ICU admission <sup>a</sup><br>n=62 | p |
| --- | --- | --- | --- | --- |
| Fevers/sweats/chills | 319 (94%) | 258 (92%) | 61 (98%) | 0.09 |
| **Headache** | **242 (71%)** | **205 (73%)** | **37 (60%)** | **0.03** |
| Anorexia/nausea/vomiting | 228 (67%) | 189 (68%) | 39 (63%) | 0.46 |
| Abdominal pain | 114 (33%) | 89 (32%) | 25 (40%) | 0.20 |
| **Diarrhoea** | **88 (26%)** | **59 (21%)** | **29 (47%)** | **<0.0001** |
| Gastrointestinal symptoms | 261 (77%) | 211 (76%) | 50 (81%) | 0.46 |
| Back pain | 55 (16%) | 41 (15%) | 14 (23%) | 0.13 |
| Lethargy | 158 (46%) | 127 (46%) | 31 (50%) | 0.52 |
| Arthralgia/myalgia | 274 (80%) | 227 (81%) | 47 (76%) | 0.32 |
| Cough | 96 (28%) | 73 (26%) | 23 (37%) | 0.08 |
| **Dyspnoea** | **35 (10%)** | **19 (7%)** | **16 (26%)** | **<0.0001** |
| **Haemoptysis (symptom)** | **17 (5%)** | **10 (4%)** | **7 (11%)** | **0.02** |

Absolute number (%) presented.
Statistically significant associations are highlighted in bold.
<sup>a</sup> Before hospital discharge from that presentation

**Supplementary Table 5.** Clinical signs at presentation in the 341 presentations in 309 episodes of leptospirosis and the association with a requirement for ICU before hospital discharge.

|  | All presentations<br>n=341 | Did not require ICU admission <sup>a</sup><br>n=279 | Required ICU admission <sup>a</sup><br>n=62 | p |
| --- | --- | --- | --- | --- |
| Conjunctival suffusion | 87 (26%) | 71 (25%) | 16 (26%) | 0.95 |
| Jaundice | 9 (3%) | 5 (2%) | 4 (6%) | 0.06 |
| **Evidence of bleeding** | **31 (9%)** | **20 (7%)** | **11 (18%)** | **0.009** |
| Rash | 26 (8%) | 19 (7%) | 7 (11%) | 0.29 |
| **Hepatomegaly** | **12 (4%)** | **7 (3%)** | **5 (8%)** | **0.048** |
| Lymphadenopathy | 22 (6%) | 16 (6%) | 6 (10%) | 0.26 |
| **Abnormal respiratory auscultation** | **61 (18%)** | **27 (10%)** | **34 (55%)** | **<0.001** |
| **Oliguria** | **60 (18%)** | **27 (10%)** | **33 (53%)** | **<0.001** |
| Temperature (°C) | 38.1 (37.2-39.0) | 38.1 (37.2-38.9) | 38.0 (37.3-39.1) | 0.94 |
| **Heart rate (beats/minute)** | **100 (89-113)** | **98 (87-110)** | **111 (92-122)** | **0.0006** |
| **Systolic blood pressure (mmHg)** | **119 (108-130)** | **120 (110-132)** | **107 (97-117)** | **0.0001** |
| **Diastolic blood pressure (mmHg)** | **70 (65-79)** | **72 (65-80)** | **66 (60-74)** | **0.0001** |
| **Mean arterial pressure (mmHg)** | **87 (79-96)** | **88 (81-97)** | **79 (73-89)** | **0.0001** |
| **Respiratory rate (breaths/minute)** | **18 (18-21)** | **18 (18-20)** | **20 (18-24)** | **0.0001** |
| Impaired consciousness | 10 (3%) | 7 (3%) | 3 (5%) | 0.40 |
| **SaO<sub>2</sub>/FiO<sub>2</sub> ratio** | **467 (457-471)** | **467 (457-471)** | **462 (451-468)** | **0.0004** |
| **Leptospirosis in initial differential diagnosis** | **216 (63%)** | **194 (70%)** | **22 (35%)** | **<0.001** |

. Statistically significant associations are highlighted in bold.
<sup>a</sup> Before hospital discharge from that presentation

**Supplementary Table 6.** Laboratory values at presentation in the 341 presentations in 309 episodes and the association with a requirement for ICU before hospital discharge.

| Laboratory values at presentation | Number with data <sup>a</sup> | All | Did not require ICU admission <sup>b</sup><br>n=279 | Required ICU admission <sup>b</sup><br>n=62 | p |
| --- | --- | --- | --- | --- | --- |
| **Haemoglobin (g/dL)** | **331** | **142 (130-150)** | **143 (134-152)** | **127 (119-147)** | **0.0001** |
| **Haematocrit** | **326** | **0.41 (0.38-0.44)** | **0.42 (0.39-0.44)** | **0.38 (0.34-0.42)** | **0.0001** |
| **White cell count (x 10<sup>9</sup>/L)** | **329** | **8.4 (6.3-11.2)** | **8.2 (6.3-10.9)** | **9.6 (7.1-12.7)** | **0.03** |
| **Neutrophil count (x 10<sup>9</sup>/L)** | **329** | **7.1 (5.2-9.6)** | **6.8 (5.0-9.1)** | **8.5 (6.3-10.9)** | **0.003** |
| **Lymphocyte count (x 10<sup>9</sup>/L)** | **329** | **0.6 (0.4-0.8)** | **0.6 (0.4-0.8)** | **0.5 (0.3-0.7)** | **0.0004** |
| **Platelet count (x 10<sup>9</sup>/L)** | **325** | **145 (105-183)** | **152 (118-188)** | **78 (49-132)** | **0.0001** |
| Sodium (mmol/L) | 330 | 134 (131-135) | 134 (131-135) | 133 (129-135) | 0.08 |
| **Potassium (mmol/L)** | **330** | **3.7 (3.5-4.0)** | **3.7 (3.5-4.0)** | **3.6 (3.3-3.9)** | **0.03** |
| **Bicarbonate (mmol/L)** | **329** | **24 (22-26)** | **24 (22-26)** | **23 (21-24)** | **0.0002** |
| **Urea (mmol/L)** | **331** | **5.4 (4.5-7.4)** | **5.1 (4.3-6.4)** | **9.0 (5.9-15.0)** | **0.0001** |
| **Creatinine (μmol/L)** | **331** | **100 (83-127)** | **98 (82-115)** | **136 (95-225)** | **0.0001** |
| **Corrected calcium (mmol/L)** | **331** | **2.25 (2.20-2.32)** | **2.26 (2.21-2.32)** | **2.23 (2.12-2.29)** | **0.005** |
| Glucose (mmol/L) | 331 | 6.3 (5.7-7.2) | 6.3 (5.7-7.1) | 6.5 (5.8-7.3) | 0.34 |
| Bilirubin (μmol/L) | 330 | 15 (10-22) | 15 (10-22) | 14 (10-20) | 0.11 |
| **Albumin (g/dL)** | **331** | **36 (32-40)** | **37 (33-40)** | **29 (27-34)** | **0.0001** |
| **Gamma-glutamyl transferase (GGT) (U/L)** | **331** | **41 (20-93)** | **39 (18-86)** | **56 (28-110)** | **0.007** |
| Serum alkaline phosphatase (SAP) (U/L) | 331 | 79 (63-119) | 78 (62-114) | 91 (67-174) | 0.06 |
| **Alanine aminotransferase (ALT) (U/L)** | **331** | **47 (28-93)** | **44 (27-89)** | **63 (34-108)** | **0.047** |
| **Aspartate Aminotransferase (AST) (U/L)** | **330** | **49 (31-94)** | **45 (30-82)** | **68 (44-136)** | **0.001** |
| Lactate dehydrogenase (U/L) | 328 | 281 (234-350) | 279 (236-345) | 288 (234-378) | 0.32 |
| **C-reactive protein (mg/L)** | **218** | **165 (100-243)** | **148 (88-220)** | **243 (157-320)** | **0.0001** |
| **Creatine kinase (U/L)** | **116** | **180 (79-804)** | **114 (42-349)** | **350 (104-1110)** | **0.0004** |
| **International normalised ratio (INR)** | **109** | **1.1 (1.1-1.2)** | **1.1 (1.0-1.2)** | **1.1 (1.1-1.3)** | **0.046** |
| **Lactate (mmol/L)** | **243** | **1.4 (1.0-2.0)** | **1.3 (0.9-1.7)** | **1.9 (1.4-2.7)** | **0.0001** |

Median (interquartile range) presented.
Statistically significant associations are highlighted in bold.
<sup>a</sup> Not all tests were performed in all patients at presentation
<sup>b</sup> Before hospital discharge from that presentation.

## References

1. Rajapakse S, Fernando N, Dreyfus A, Smith C, Rodrigo C. Leptospirosis. Nature Reviews Disease Primers. 2025;11(1):32. doi. 10.1038/s41572-025-00614-5.

2. Costa F, Hagan JE, Calcagno J, Kane M, Torgerson P, Martinez-Silveira MS, et al. Global Morbidity and Mortality of Leptospirosis: A Systematic Review. PLoS Negl Trop Dis. 2015;9(9):e0003898. doi. 10.1371/journal.pntd.0003898.

3. Torgerson PR, Hagan JE, Costa F, Calcagno J, Kane M, Martinez-Silveira MS, et al. Global Burden of Leptospirosis: Estimated in Terms of Disability Adjusted Life Years. PLoS Negl Trop Dis. 2015;9(10):e0004122. doi. 10.1371/journal.pntd.0004122.

4. Chawla V, Trivedi TH, Yeolekar ME. Epidemic of leptospirosis: an ICU experience. J Assoc Physicians India. 2004;52:619–22.

5. Miailhe AF, Mercier E, Maamar A, Lacherade JC, Le Thuaut A, Gaultier A, et al. Severe leptospirosis in non-tropical areas: a nationwide, multicentre, retrospective study in French ICUs. Intensive Care Med. 2019;45(12):1763–73. doi. 10.1007/s00134-019-05808-6.

6. Delmas B, Jabot J, Chanareille P, Ferdynus C, Allyn J, Allou N, et al. Leptospirosis in ICU: A Retrospective Study of 134 Consecutive Admissions. Crit Care Med. 2018;46(1):93–9. doi. 10.1097/ccm.0000000000002825.

7. Hart S, Smith S, Nash B, Stratton H, Rosengren P, Goud RS, et al. The management and clinical course of patients admitted to the intensive care unit with leptospirosis in a referral hospital in Far North Queensland, Tropical Australia. Acta Trop. 2026;279:108154. doi. 10.1016/j.actatropica.2026.108154.

8. Smith S, Hanson J. Improving the mortality of severe leptospirosis. Intensive Care Medicine. 2020;46(4):827–8. doi. 10.1007/s00134-020-05925-7.

9. Smith S, Kennedy BJ, Dermedgoglou A, Poulgrain SS, Paavola MP, Minto TL, et al. A simple score to predict severe leptospirosis. PLoS Negl Trop Dis. 2019;13(2):e0007205. doi. 10.1371/journal.pntd.0007205.

10. Galdino GS, de Sandes-Freitas TV, de Andrade LGM, Adamian CMC, Meneses GC, da Silva Junior GB, et al. Development and validation of a simple machine learning tool to predict mortality in leptospirosis. Scientific Reports. 2023;13(1):4506. doi. 10.1038/s41598-023-31707-4.

11. Haake DA, Levett PN. Leptospirosis in humans. Curr Top Microbiol Immunol. 2015;387:65–97. doi. 10.1007/978-3-662-45059-8_5.

12. Izurieta R, Galwankar S, Clem A. Leptospirosis: The “mysterious” mimic. J Emerg Trauma Shock. 2008;1(1):21–33. doi. 10.4103/0974-2700.40573.

13. Stratton H, Pownell C, Sandeman M, Rosengren P, Price C, Stewart AGA, et al. Deciphering the “Doxycycline Deficiency Syndrome”: Defining the Characteristic Clinical, Laboratory and Imaging Findings in Patients Presenting with Leptospirosis, Q Fever and Rickettsial Diseases in Far North Queensland, Tropical Australia. Preprints: Preprints; 2026. doi:10.20944/preprints202606.0044.v1.

14. National Early Warning Score (NEWS) 2: Standardising the assessment of acute-illness severity in the NHS. London: Royal College of Physicians; 2017.

15. Vincent JL, Moreno R, Takala J, Willatts S, De Mendonça A, Bruining H, et al. The SOFA (Sepsis-related Organ Failure Assessment) score to describe organ dysfunction/failure. On behalf of the Working Group on Sepsis-Related Problems of the European Society of Intensive Care Medicine. Intensive Care Med. 1996;22(7):707–10. doi. 10.1007/bf01709751.

16. Rosengren P, Johnston L, Ismail I, Smith S, Hanson J. The Characteristics of Patients That Develop Severe Leptospirosis: A Scoping Review. Pathogens. 2025;14(12). doi. 10.3390/pathogens14121268.

17. National Notifiable Disease Surveillance System Canberra: Australian Government. Department of Health Disability and Aging; 2026 [Available from: https://www.cdc.gov.au/diseases/surveillance-systems-and-networks/national-notifiable-diseasessurveillance-system-nndss.

18. Stratton H, Rosengren P, Kinneally T, Prideaux L, Smith S, Hanson J. Presentation and Clinical Course of Leptospirosis in a Referral Hospital in Far North Queensland, Tropical Australia. Pathogens. 2025;14(7):643.

19. Lee N, Tanaka T, Koizumi N, Coughlan C, Sayo AR, Ariyoshi K, et al. Leptospirosis-clinical review and updates on therapeutics. Lancet Infect Dis. 2026. doi. 10.1016/s1473-3099(26)00060-5.

20. Franklin RC, King JC, Aitken PJ, Elcock MS, Lawton L, Robertson A, et al. Aeromedical retrievals in Queensland: A five-year review. Emerg Med Australas. 2021;33(1):34–44. doi. 10.1111/1742-6723.13559.

21. Hanson J, Smith S, Brooks J, Groch T, Sivalingam S, Curnow V, et al. The applicability of commonly used predictive scoring systems in Indigenous Australians with sepsis: An observational study. PLoS One. 2020;15(7):e0236339. doi. 10.1371/journal.pone.0236339.

22. Fox H, Hempenstall A, Pilot P, Callander E, Smith S, McDonald MI, et al. Significant healthcare resource utilisation in the management of skin and soft tissue infections in the Torres Strait, Australia. Rural Remote Health. 2024;24(2):8572. doi. 10.22605/rrh8572.

23. Queensland regions data. Queensland Government. Department of Primary Industries. Brisbane. https://www.dpi.qld.gov.au/news-media/campaigns/data-farm/regions2026 13 May 2026. Available from: https://www.dpi.qld.gov.au/news-media/campaigns/data-farm/regions.

24. Fairhead LJ, Smith S, Sim BZ, Stewart AGA, Stewart JD, Binotto E, et al. The seasonality of infections in tropical Far North Queensland, Australia: A 21-year retrospective evaluation of the seasonal patterns of six endemic pathogens. PLOS Glob Public Health. 2022;2(5):e0000506. doi. 10.1371/journal.pgph.0000506.

25. Leptospirosis Reference Laboratory: Queensland Health. Queensland Government.; 2026 [Available from: https://www.health.qld.gov.au/public-health/forensic-and-scientific-services/testing-analysis/disease-investigation-and-analysis/leptospirosis-reference-laboratory.

26. Wright SW, Hantrakun V, Rudd KE, Lau CY, Lie KC, Chau NVV, et al. Enhanced bedside mortality prediction combining point-of-care lactate and the quick Sequential Organ Failure Assessment (qSOFA) score in patients hospitalised with suspected infection in southeast Asia: a cohort study. Lancet Glob Health. 2022;10(9):e1281–e8. doi. 10.1016/s2214-109x(22)00277-7.

27. Seymour CW, Liu VX, Iwashyna TJ, Brunkhorst FM, Rea TD, Scherag A, et al. Assessment of Clinical Criteria for Sepsis: For the Third International Consensus Definitions for Sepsis and Septic Shock (Sepsis-3). Jama. 2016;315(8):762–74. doi. 10.1001/jama.2016.0288.

28. Redfern OC, Smith GB, Prytherch DR, Meredith P, Inada-Kim M, Schmidt PE. A Comparison of the Quick Sequential (Sepsis-Related) Organ Failure Assessment Score and the National Early Warning Score in Non-ICU Patients With/Without Infection. Crit Care Med. 2018;46(12):1923–33. doi. 10.1097/ccm.0000000000003359.

29. Bone RC, Balk RA, Cerra FB, Dellinger RP, Fein AM, Knaus WA, et al. Definitions for sepsis and organ failure and guidelines for the use of innovative therapies in sepsis. The ACCP/SCCM Consensus Conference Committee. American College of Chest Physicians/Society of Critical Care Medicine. Chest. 1992;101(6):1644–55. doi. 10.1378/chest.101.6.1644.

30. Moore CC, Hazard R, Saulters KJ, Ainsworth J, Adakun SA, Amir A, et al. Derivation and validation of a universal vital assessment (UVA) score: a tool for predicting mortality in adult hospitalised patients in sub-Saharan Africa. BMJ Global Health. 2017;2(2):e000344. doi. 10.1136/bmjgh-2017-000344.

31. Pandharipande PP, Shintani AK, Hagerman HE, St Jacques PJ, Rice TW, Sanders NW, et al. Derivation and validation of Spo2/Fio2 ratio to impute for Pao2/Fio2 ratio in the respiratory component of the Sequential Organ Failure Assessment score. Crit Care Med. 2009;37(4):1317–21. doi. 10.1097/CCM.0b013e31819cefa9.

32. Marotto PC, Nascimento CM, Eluf-Neto J, Marotto MS, Andrade L, Sztajnbok J, et al. Acute lung injury in leptospirosis: clinical and laboratory features, outcome, and factors associated with mortality. Clin Infect Dis. 1999;29(6):1561–3. doi. 10.1086/313501.

33. Andrade L, Cleto S, Seguro AC. Door-to-dialysis time and daily hemodialysis in patients with leptospirosis: impact on mortality. Clin J Am Soc Nephrol. 2007;2(4):739–44. doi. 10.2215/cjn.00680207.

34. Karnik ND, Patankar AS. Leptospirosis in Intensive Care Unit. Indian J Crit Care Med. 2021;25(Suppl 2):S134–s7. doi. 10.5005/jp-journals-10071-23852.

35. Rajapakse S, Rodrigo C, Haniffa R. Developing a clinically relevant classification to predict mortality in severe leptospirosis. J Emerg Trauma Shock. 2010;3(3):213–9. doi. 10.4103/0974-2700.66519.

36. Villar J, Short JH, Lighthall G. Lactate Predicts Both Short-and Long-Term Mortality in Patients With and Without Sepsis. Infect Dis (Auckl). 2019;12:1178633719862776. doi. 10.1177/1178633719862776.

37. Ihalainen V, Pölkki A, Moser A, Takala J, Jakob SM, Bendel S, et al. The Glasgow Coma Scale as a predictor of mortality in intensive care: relevance of underlying cause of impaired consciousness. J Crit Care. 2026;94:155605. doi. 10.1016/j.jcrc.2026.155605.

38. Colunga-Lozano LE, Foroutan F, Rayner D, De Luca C, Hernández-Wolters B, Couban R, et al. Clinical judgment shows similar and sometimes superior discrimination compared to prognostic clinical prediction models: a systematic review. Journal of Clinical Epidemiology. 2024;165:111200. doi. 10.1016/j.jclinepi.2023.10.016.

39. Ko AI, Reis MG, Dourado CMR, Johnson WD, Jr., Riley LW. Urban epidemic of severe leptospirosis in Brazil. The Lancet. 1999;354(9181):820–5. doi. 10.1016/S0140-6736(99)80012-9.

40. Katz AR, Ansdell VE, Effler PV, Middleton CR, Sasaki DM. Assessment of the clinical presentation and treatment of 353 cases of laboratory-confirmed leptospirosis in Hawaii, 1974-1998. Clin Infect Dis. 2001;33(11):1834–41. doi. 10.1086/324084.

41. Berman SJ, Tsai CC, Holmes K, Fresh JW, Watten RH. Sporadic anicteric leptospirosis in South Vietnam. A study in 150 patients. Ann Intern Med. 1973;79(2):167–73. doi. 10.7326/0003-4819-79-2-167.

42. Levin N, Horton D, Sanford M, Horne B, Saseendran M, Graves K, et al. Failure of vital sign normalization is more strongly associated than single measures with mortality and outcomes. The American Journal of Emergency Medicine. 2020;38(12):2516–23. doi. 10.1016/j.ajem.2019.12.024.

43. Jirawannaporn S, Limothai U, Tachaboon S, Dinhuzen J, Kiatamornrak P, Chaisuriyong W, et al. Rapid and sensitive point-of-care detection of Leptospira by RPA-CRISPR/Cas12a targeting lipL32. PLoS Negl Trop Dis. 2022;16(1):e0010112. doi. 10.1371/journal.pntd.0010112.

